# Biomarkers of protection against controlled human SARS-CoV-2 Delta variant breakthrough infection

**DOI:** 10.64898/2026.07.24.26358855

**Authors:** Anika Singanayagam, Helen R. Wagstaffe, Lydia Slater, Polly Fox-Sheehan, Meng-San Wu, Andrew Mawer, Hannah Preston-Jones, Orlagh Daly, Jen Mae Low, Raquel Lopez Ramon, Eileen Hughes, Jie Zhou, Anjna Badhan, Jon Guy, Stephanie A. Harris, Samuel P. Smith, Melissa Govender, Stephen M. Laidlaw, Tom Tipton, Iman Satti, Merenienla Yaden, Stephanie Ascough, Ksenia Sukhova, Maya Moshe, Joanne McKenzie, Henna Siddiqui, Alberta Ateere, Beatrice Nassanga, Freya Stiff, David S. Khoury, Arnold Reynaldi, Miles P. Davenport, Miles Carroll, Ryan S. Thwaites, Graham P. Taylor, Wendy Barclay, Margherita Bracchi, Helen McShane, Christopher Chiu, the COVHIC002 study team

## Abstract

**Background:** Improved understanding of how variants cause breakthrough infection, despite pre-existing immunity, is needed to advance development of next-generation SARS-CoV-2 vaccines, including those that may provide cross-variant protection or block transmission. SARS-CoV-2 controlled human infection models (CHIMs) may therefore identify correlates of protection and accelerate the development of new interventions.

**Methods:** Healthy vaccinated adults aged 18-30 years were inoculated intranasally in a stepwise dose-escalation CHIM with doses from 1x10^2^ TCID_50_ to 1x10^6^ TCID_50_ of SARS-CoV-2 Delta variant. Within the 1x10^6^ TCID_50_ group, participants were selected for serum neutralising antibody titres (NT_50_) ≤1:80. Post-inoculation, participants were quarantined for up to 14 days. Outpatient follow-up continued for 12 months. The primary aim was to elicit safe, well-tolerated Delta SARS-CoV-2 breakthrough infection at a rate of over 50%.

**Findings:** Forty-six participants were inoculated; 22 during dose-escalation with no resultant infections, and 24 at the highest dose, of whom 18 were screened for low serum neutralising antibodies. Sustained infection with mild-to-moderate symptoms occurred in 33% (6/18) of the sero- selected group, with highly variable viral loads, viral emissions and symptoms. Serum neutralising antibody, anti-N IgG and to a lesser extent, mucosal anti-S IgA and N-specific T cells most strongly predicted protection from virologically-defined infection. Higher neutralisation, serum and nasal anti-N IgG level, and N-specific T cell responses correlated with lower viral load, while baseline nasal anti-S IgA was associated with lower symptom scores. Transient infection was additionally observed in 6 participants and was associated with higher baseline N-specific T cell responses than those that developed sustained infection.

**Interpretation:** Susceptibility to SARS-CoV-2 breakthrough infection in those with hybrid immunity is strongly associated with low levels of pre-existing antibody, but the diversity of immune markers associated with protection implies that additional benefits may be conferred by multi-pronged immunity.

**Funding:** Wellcome Trust

**Research in context:** *Evidence before this study:* To identify other published SARS-CoV-2 controlled human infection models (CHIM), a search on PubMed was carried out on 4th March 2026. The search terms used were ((“controlled human infection”) OR (“human challenge”)) and ((SARS-CoV-2) OR (COVID-19)) and (“clinical trial”). Two clinical studies were identified. The first study involved healthy adult 18-29 year olds, seronegative to SARS-CoV-2 inoculated with 1x101 TCID50 pre-Alpha (wild-type) SARS-CoV-2. Eighteen of 34 (53%) participants became infected. The model was safe and well-tolerated and there were no study-related serious adverse events. Mild to moderate symptoms were reported by 16 of 18 (89%) infected individuals while 2 had virtually no symptoms. 14 of 18 (78%) of participants reported smell disturbance measured by the University of Pennsylvania Smell Identification Test (UPSIT). Viral detection by qPCR became quantifiable in throat swabs from 40 hours (∼1.67 days) post-inoculation and nose swabs at 58 hours (∼2.4 days). Viral load peaked in the throat at 112 hours (∼4.2 days) post-inoculation and later at 148 hours (∼6.2 days) post-inoculation in the nose. The second study involved healthy adult 18-30 year olds, seropositive to SARS-CoV-2 inoculated with escalating doses (1x10^1^-1x10^5^ TCID_50_) of the same pre-Alpha variant. Thirty-six participants were inoculated and no sustained infection meeting the protocol-defined definition of infection was induced. Five (14%) of 36 volunteers were considered to have transient (brief) infections, based on the kinetic of their PCR-positive swabs. In the first study, functionally-complete protection was associated with early increases in innate and adaptive cell abundance in the nose post-inoculation, higher pre-existing chemokine levels (particularly CCL13) in the nasal lining fluid and cross-reactive T cell responses. Transient infections (PCR positivity outside of residual inoculum not meeting the protocol-defined criteria for infection) were present in both studies. In the second study, transient infection was associated with significantly lower baseline mucosal and systemic SARS-CoV- 2 antibody levels and significantly lower peripheral IFN-γ producing CD8^+^ T-cell against a SARS-CoV-2 peptide pool than uninfected participants.

*Added value of this study:* With near-universal seropositivity to SARS-CoV-2, understanding the factors that influence how variants cause breakthrough infection is critical. The optimisation of a model that can induce safe and tolerable infection in a large proportion of participants is necessary for SARS-CoV-2 CHIMs to be used as a tool for next-generation vaccine development. Our study is the first SARS-CoV-2 CHIM of seropositive individuals to induce sustained, protocol defined infection, albeit with an infection rate of 33%. Despite the low number of infected individuals, we identified several potential correlates of protection against breakthrough Delta SARS-CoV-2 infection beyond serum neutralising antibodies.

*Implications of all the available evidence:* This study establishes the framework for SARS-CoV-2 CHIM conduct, using sero-selection to increase attack rate in the same way it is used for influenza human challenge studies, and a process for defining quantitative correlates of protection. Further work will optimise model parameters with Omicron subvariants to result in infection rates of ≥50% to support testing of novel interventions.

## Introduction

Available SARS-CoV-2 vaccines are highly effective in reducing severe disease, yet their capacity to prevent upper respiratory infection is limited. Vaccines able to induce functionally- sterilising immunity could have major impacts on transmission reduction, conferring secondary protection for vulnerable groups. However, to advance development of next-generation vaccines, improved understanding of how breakthrough SARS-CoV-2 infection occurs despite pre-existing immunity is needed. Neutralising antibody has previously been shown to correlate with protection against symptomatic SARS-CoV-2 disease^1^ but protection against infection itself is less well understood. Furthermore, hybrid immunity (gained from the combination of infection and vaccination) affords better protection than vaccination or infection alone^2^. Potential contributory mechanisms include a more robust and durable immune response with higher levels of neutralising antibodies, more robust and polyfunctional T cell responses^3^ and mucosal immunity^4^, but their relative importance is unclear.

SARS-CoV-2 controlled human infection models (CHIMs) can uniquely address these outstanding questions and accelerate development of new interventions by defining correlates of protection and efficacy testing early during clinical development. The strength of these studies lies in their controlled nature, with the use of standardised amounts of well-characterised virus enabling the exact longitudinal measurement of viral kinetics^5^, transmission dynamics^6^, immune responses and associations with protection and clearance of infection^5,7^.

We previously established a safe and acceptable SARS-CoV-2 human challenge model with the wild-type pre-Alpha virus in seronegative, unvaccinated volunteers^8^. However, challenge with vaccine homologous pre-Alpha virus in vaccinated volunteers did not result in sustained infection^4^. In 2021, the Delta variant (B.617.1) emerged and caused a surge in natural breakthrough infections^9^. Therefore, to support further study of next-generation vaccines with cross-variant and transmission-blocking capabilities, we set out to develop a breakthrough challenge model using Delta SARS-CoV-2. The primary aim was to identify model parameters that would allow safe, well-tolerated breakthrough infection with a ≥50% attack rate.

Here, we show that mild-to-moderate infection can be safely induced under controlled conditions with Delta SARS-CoV-2. Breakthrough infection manifested variable viral loads, viral emissions and symptoms, demonstrating a heterogeneity of infection outcomes in healthy adults with pre-existing immunity. Thus, we were able to identify multiple immune measures that predicted breakthrough infection.

## Methods

### Ethics Statement

This study was approved by the UK Health Research Authority Specialist Ad-Hoc Research Ethics Committee (Ref. 22/UK/0001; IRAS ID: 318173). Written informed consent was obtained from all participants prior to screening and study enrolment. The study was overseen by a Trial Steering Committee with advice from an independent Data and Safety Monitoring Committee and was registered with the ISRCTN Registry (ISRCTN94747181).

### Study design

Healthy adults aged 18-30 years who had completed a full course of SARS-CoV-2 vaccination, with or without prior infection, were recruited into this multi-centre, phase 1, open-label study. The study was carried out in London and Oxford, UK from November 2022 to June 2025. See Supplementary Appendix for study protocol detailing screening, eligibility and recruitment procedures and for detailed methods.

The primary aim was to identify a safe, well-tolerated dose of SARS-CoV-2 Delta variant (B.1.617.2) that resulted in an infection rate of ≥50% when inoculating healthy vaccinated volunteers. Participants were inoculated with 10^2^-10^6^ 50% tissue culture infectious dose (TCID_50_) SARS-CoV-2 Delta variant in a dose escalation protocol. The study timeline detailing sampling procedures is shown in Fig. 1a.

**Fig. 1.**
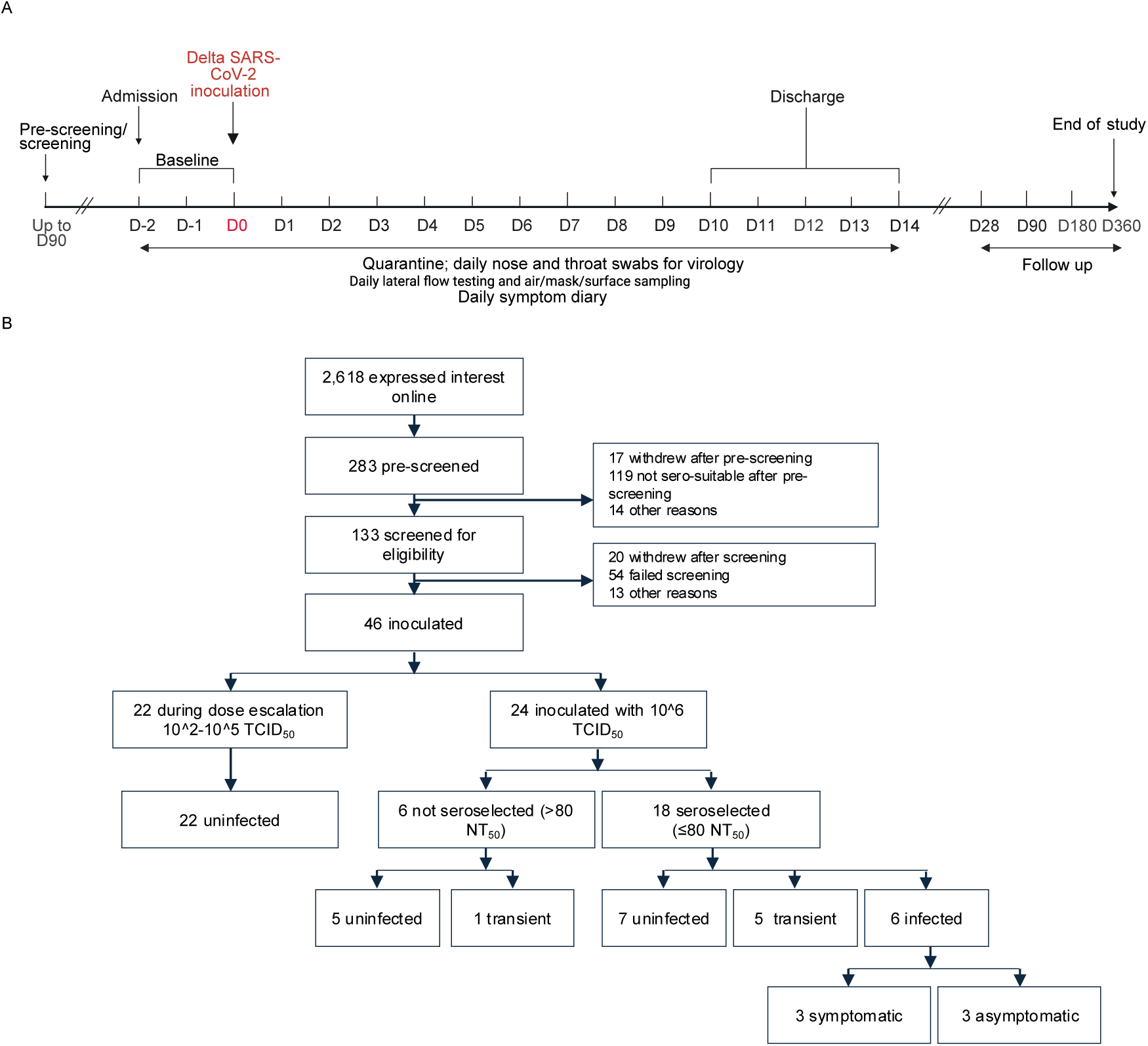
Study timeline and CONSORT. **a**, Study timeline showing inoculation and sample collection timepoints. **b**, Consort diagram showing participants pre-screened, enrolled and inoculated, with outcomes.

### Clinical assessments

Safety was assessed with blood tests, spirometry, ECG and clinical assessments (vital signs, symptom diaries, clinical examination). Prior to inoculation, multiplex respiratory pathogen PCR was carried out to exclude co-existing infection. Symptom scores were recorded by participants in symptom diaries.

### Procedures

PBMC, serum and nasosorption samples for immunology, mid-turbinate nose and throat swabs for viral load detection and air, surface and face mask sampling for viral emissions were collected at regular intervals during a 14 day quarantine period before and after inoculation on day 0 (Fig. 1a). Pre-screening visits for serology testing was up to 90 days prior to inoculation.

### Virological assessments

Nose and throat swabs were subjected to virological analyses using RT-qPCR for the envelope (E) and nucleocapsid (N) gene, (with human RNAse P as process control), plaque assay, and lateral flow antigen test (LFT) (Flowflex). Mask, air and environmental samples were subjected to RT-qPCR for the E gene, virus culture, and were also quantified for human housekeeping gene 18S rRNA by PCR (see Supplementary Appendix for further detail).

### Immunological assessments

For participant seroselection, SARS-CoV-2 specific antibodies were measured at a pre-screening visit by live virus neutralisation assay and anti-spike (S) and anti-N binding IgG was measured by Roche Electrochemiluminescence (ECL) assay. Baseline (day -2, -1 or day 0 pre-inoculation) and post-inoculation timepoint antibody assessments were completed retrospectively on frozen samples after study completion by microneutralisation assay (MNA) or Meso Scale Discovery (MSD) assay.

*Ex vivo* IFN-g T cell ELISpot assays were performed on freshly isolated peripheral blood mononuclear cells (PBMC) as previously described^4^. For 3-colour fluorospot, cryopreserved PBMC were thawed and CTL (ImmunoSpot) fluorospot kit was run following the manufacturer instructions, as previously described^10^.

### Statistical analysis

Statistical analysis was performed using GraphPad Prism version 10.4.1 and R version 4.5.1. correlation plots were generated using corrplot. Statistical tests used are indicated in the figure legends. A p value of less than 0.05 was used to indicate significance. See Supplementary Appendix for univariate analysis detail.

## Results

### High inoculum dose and low pre-existing immunity are necessary for Delta breakthrough infection

Forty-six healthy adult volunteers were inoculated with SARS-CoV-2 Delta variant. Twenty-two were inoculated during the dose escalation phase (1x10^2^ (n=5), 1x10^3^ (n=5), 1x10^4^ (n=6), 1x10^5^ (n=6)) with no resulting infections. Twenty-four were inoculated at the highest dose of 1x10^6^ TCID_50_. After 6 participants remained uninfected at the highest dose, participants were pre-selected using serum live virus neutralising antibody titre (NT_50_) ≤1:80. Eighteen seroscreened participants were subsequently challenged, and 6 met the protocol-defined criteria for infection (an infection rate of 33%). In addition, six transient infections were also observed, similar to the previous pre-Alpha SARS-CoV-2 controlled human infection studies, 1 in the non-seroselected group and 5 in the seroselected group^4,7^. The remaining 12 participants were deemed uninfected (Fig. 1b).

Amongst those inoculated with 1x10^6^ TCID_50_, mean age was 25 (18-29) years and 58% were male. There were no demographic factors significantly associated with infection outcome (Table 1). All participants had previously received a full course of SARS-CoV-2 vaccine. 83% reported at least one previous SARS-CoV-2 infection, with a lower proportion in the sustained infection group compared with the transient and uninfected groups (67%, 83%, 83% respectively) (Table 1).

**Table 1:** Baseline demographics, vaccination and infection history, symptoms and adverse events of the 1x10^6^ TCID_50_ inoculated group. P value calculated with Mann-Whitney test or Fisher’s exact test, p=<0.05 = significant.

|  | **Total**<br>**($1 \times 10^6$**<br>**TCID<sub>50</sub>**<br>**dose)** | **Infected** | **Transient** | **Uninfected** | **Sero-selected** | **Non sero-**<br>**selected** | **P value**<br>**(infected vs**<br>**combined**<br>**transient plus**<br>**uninfected**<br>**group)** |
| --- | --- | --- | --- | --- | --- | --- | --- |
| **Characteristic** | **n=24** | **n=6** | **n=6** | **n=12** | **n=18** | **n=6** | **n=6 vs n=18** |
| Age (years) |  |  |  |  |  |  | 0.886 |
| Mean (s.d) | 25.2 (3.8) | 24.7 (4.1) | 25.7 (4.2) | 25.3 (3.8) | 24.8 (3.9) | 26.5 (3.3) |  |
| Min, Max | 18, 29 | 19, 29 | 18, 29 | 19, 29 | 18, 29 | 19, 29 |  |
| Gender, n (%) |  |  |  |  |  |  | 0.192 |
| Male | 14 (58) | 2 (33) | 4 (67) | 8 (67) | 11 (61) | 3 (50) |  |
| Female | 10 (42) | 4 (67) | 2 (33) | 4 (33) | 7 (39) | 3 (50) |  |
| Race, n (%) |  |  |  |  |  |  | 0.0785 |
| White | 19 (79) | 3 (50) | 5 (83) | 11 (92) | 14 (78) | 5 (83) |  |
| Mixed ethnicity | 4 (17) | 3 (50)<br>2 White/Asian<br>1 White/Asian<br>Indian | 0 | 1 (8)<br>White/Black<br>Caribbean | 3 (17) | 1 (17) |  |
| Asian or Asian British | 1 (4) | 0 | 1 (17) | 0 | 1 (6) | 0 |  |
| BMI (Kg m<sup>-2</sup>) |  |  |  |  |  |  | 0.431 |
| Mean (range, s.d) | 23.86 (18-29, 3.9) | 23.05 (19-29,<br>4.1) | 23.69 (18-29, 4.2) | 24.35 (19-29,<br>3.9) | 23.55 (18-29,<br>4.0) | 24.80 (21-29,<br>3.3) |  |
| Vaccination history |  |  |  |  |  |  | 0.629 |
| 3rd vaccine dose, n (%) | 16 (67) | 5 (83) | 4 (67) | 7 (58) | 11 (61) | 5 (83) |  |
| 4th vaccine dose, n (%) | 0 | 0 | 0 | 1 (8) | 0 | 1 (17) |  |
| Infection history |  |  |  |  |  |  | 0.568 |
| Previous LFT or PCR positive, n (%) | 20 (83) | 4 (67) | 5 (83) | 10 (83) | 15 (83) | 5 (83) |  |
| Antibody level at screening (up to 90 days prior to inoculation) |  |  |  |  |  |  |  |
| Anti-Spike, BAU/ml, median (IQR) | 4812 (17708) | 247 (3685) | 3112 (6188) | 6210 (16,419) | 3112 (7329) | 8537 (11,581) |  |
| Anti-Nucleocapsid, COI, median (IQR) | 19.7 (320.9) | 0.52 (30.4) | 14.0 (319.3) | 51.7 (305.7) | 4.2 (139.9) | 202.0 (320.0) |  |
| Neutralising antibody titre, NT50, median (IQR) | 40 (310) | 20 (20) | 47 (87) | 80 (310) | 40 (70) | 160 (213) |  |
| Symptoms |  |  |  |  |  |  |  |
| Any report of symptoms on 2 consecutive days, n (%) | 13 (54) | 3 (50) | 4 (67) | 6 (50) | 10 (56) | 3 (50) |  |
| Fever |  |  |  |  |  |  |  |
| >37.8°C, n (%) | 0 | 0 | 0 | 0 | 0 | 0 |  |
| Adverse events (AE) |  |  |  |  |  |  |  |
| Any serious AE | 0 | 0 | 0 | 0 | 0 |  |  |
| Clinically significant adverse events thought to be probably associated with challenge viral infection |  |  |  |  |  |  |  |
| Smell disturbance (UPSIT) (%) | 1 (4) | 1 (4) | 0 | 0 | 1 (6) | 0 |  |
| Low white cell count <2.0 × 10<sup>9</sup> per l, n | 0 | 0 | 0 | 0 | 0 | 0 |  |
| Low lymphocytes (<0.75 × 10<sup>9</sup> per l) | 0 | 0 | 0 | 0 | 0 | 0 |  |
| Low neutrophils (<1.0 × 10<sup>9</sup> per l) | 0 | 0 | 0 | 0 | 0 | 0 |  |
| Low neutrophils (<0.5 × 10<sup>9</sup> per l) | 0 | 0 | 0 | 0 | 0 | 0 |  |

There were no serious adverse events. Adverse events related to SARS-CoV-2 inoculation were mild or moderate, of short duration, and there were no symptoms or signs of post-COVID-19 condition (“long COVID”). There were no clinically significant changes in any other clinical assessments related to SARS-CoV-2 inoculation or infection. Further details on adverse events are in Table 1 and Supplementary Table 1.

### Heterogeneity of viral loads, viral emissions and symptom scores in breakthrough infection

Upper respiratory tract (URT) viral kinetics were highly varied between participants with breakthrough infection (Fig. 2a, b). Peak viral load (VL) was significantly higher in nose swabs than throat swabs (6.3 log_10_ copies/ml versus 3.1 log_10_ copies/ml, p=0.0048) (Supplementary Fig. 1a, b). PCR positive detections in transiently infected participants were primarily at earlier timepoints, day 2 and 3 post-inoculation (p.i), suggesting rapid termination of early viral replication before disease onset. Viral RNA on day 1 p.i, potentially due to residual inoculum, was detected in 18 of 24 participants in the 1x10^6^ TCID_50_ dose group (including in 7/12 uninfected participants) (Fig. 2a, b and Supplementary Fig. 1a).

**Fig. 2.**
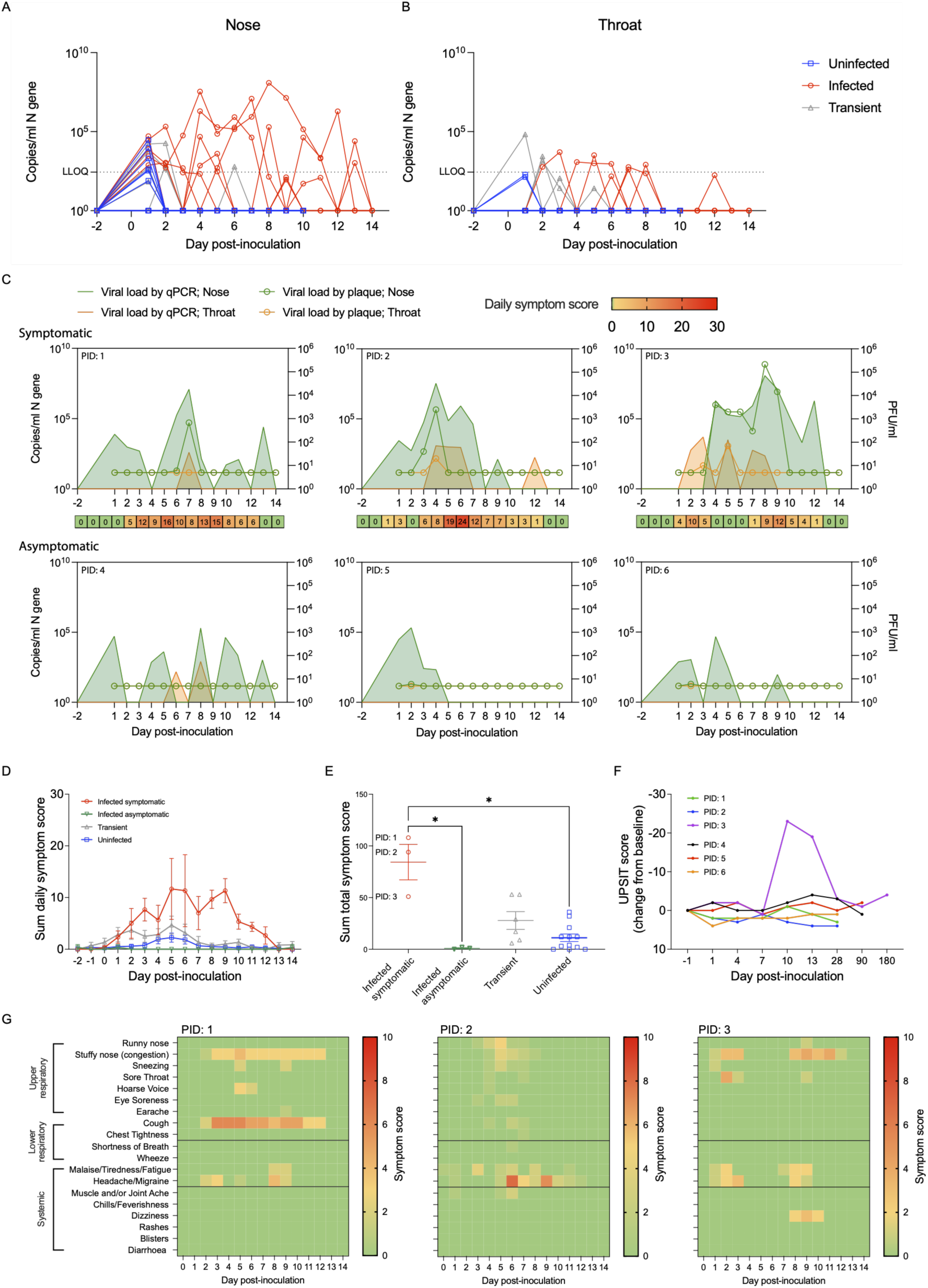
Upper respiratory tract viral loads and symptom scores are heterogeneous in breakthrough infection. Copies/ml SARS-CoV-2 N gene in swabs of the **a**, nose and **b**, throat of the three outcome groups (uninfected n=12, transient n=6 and infected n=6) measured by qPCR. Line at N gene LLOQ = 275 copies/ml. **c**, Viral load in the nose and throat measured by qPCR (left axis) and plaque assay (right axis) of the 3 symptomatic and 3 asymptomatic infected participants with a heat map of daily symptom score for the symptomatic group. **d**, Daily symptom score and **e**, sum total symptom score of the 3 outcome groups with symptomatic infected separated from the asymptomatic infected group. **f**, University of Pennsylvania smell identification test (UPSIT) score of the 6 infected participants. **g**, Symptoms experienced by the three infected symptomatic patients. Two-Way ANOVA mixed effects model with Geisser-Greenhouse correction and Tukey’s multiple comparisons test (a, b, d) or One-Way ANOVA Kruskal-Wallis test with Dunn’s multiple comparisons test (e). Mean and SEM or all points shown. *p=<0.05.

Of the 6 infected participants, 3 were symptomatic and 3 were asymptomatic. Symptomatically infected participants trended toward higher peak nasal VL (mean N gene 7.6 Log_10_ copies/ml vs 5.1 Log_10_ copies/ml, p=0.1) and higher nasal VL AUC (N gene 40.5 vs 27.6, p=0.7) than asymptomatic individuals (Fig. 2c and Supplementary Fig. 1c). Symptomatic participants also had more culturable virus detections in the URT (1-6 days detection vs 0 days detection) (Fig. 2c). Little virus was detected by plaque assay in asymptomatic or transiently infected participants suggesting a low likelihood of transmissibility (single day detections in 3 participants only) (Supplementary Fig. 1a).

Symptoms reported by the 3 symptomatically-infected participants peaked on days 5 or 9 and were mild-to-moderate in severity (Fig. 2d). Stuffy nose was the most frequently reported symptom and systemic symptoms (such as malaise, tiredness, fatigue and headache) were also common (Fig. 2f). There was no correlation between total symptom score and VL in the infected group (Supplementary Fig. 1d). Four of 6 transiently-infected participants reported symptoms on 2 or more consecutive days; these were upper respiratory and systemic in nature (Supplementary Fig. 1e). Total symptom scores were highest in the symptomatically infected (mean 84.33 vs infected asymptomatic; 1, transient; 27.83 and uninfected; 11.17) (Fig. 2e). One individual, who had the highest AUC and peak nasal VL, displayed biphasic symptomatology; symptoms reported on days 1-3, which resolved, then reappeared on day 8 (Fig. 2f). This individual also reported transient change in smell and taste between days 8 and 14, reflected in a decreased UPSIT score (−23 from baseline), which returned to baseline by day 28 (Fig. 2g). No other participants in the study reported any change in smell or taste or any prolonged symptomatology.

To estimate transmission potential of breakthrough infections, virus emitted into the air and environment was collected, as previously described^6^. In exhaled breath from face-mask samples (FMS), virus was detected from 5 out of 6 infected participants (median 2.45 Log_10_ E gene copies per sample, 95% CI 2.05-3.21). Total virus emissions detected in FMS in infected individuals correlated with the total VL detected in the nose (AUC of log_10_ VL; p=0.072, Spearman) and in the throat (p=0.022). Symptomatic infected participants had more positive viral detections in FMS, hand and surface swabs than asymptomatic infected participants (median of 5 vs 2, p=0.3). Greatest contamination of environmental surfaces and hands was by the two symptomatic participants with highest URT viral shedding. Virus was detected in air samples from only 2 participants at 1 timepoint each (both asymptomatic participants, day 3 and day 1 p.i) (Fig. 3a).

**Fig. 3.**
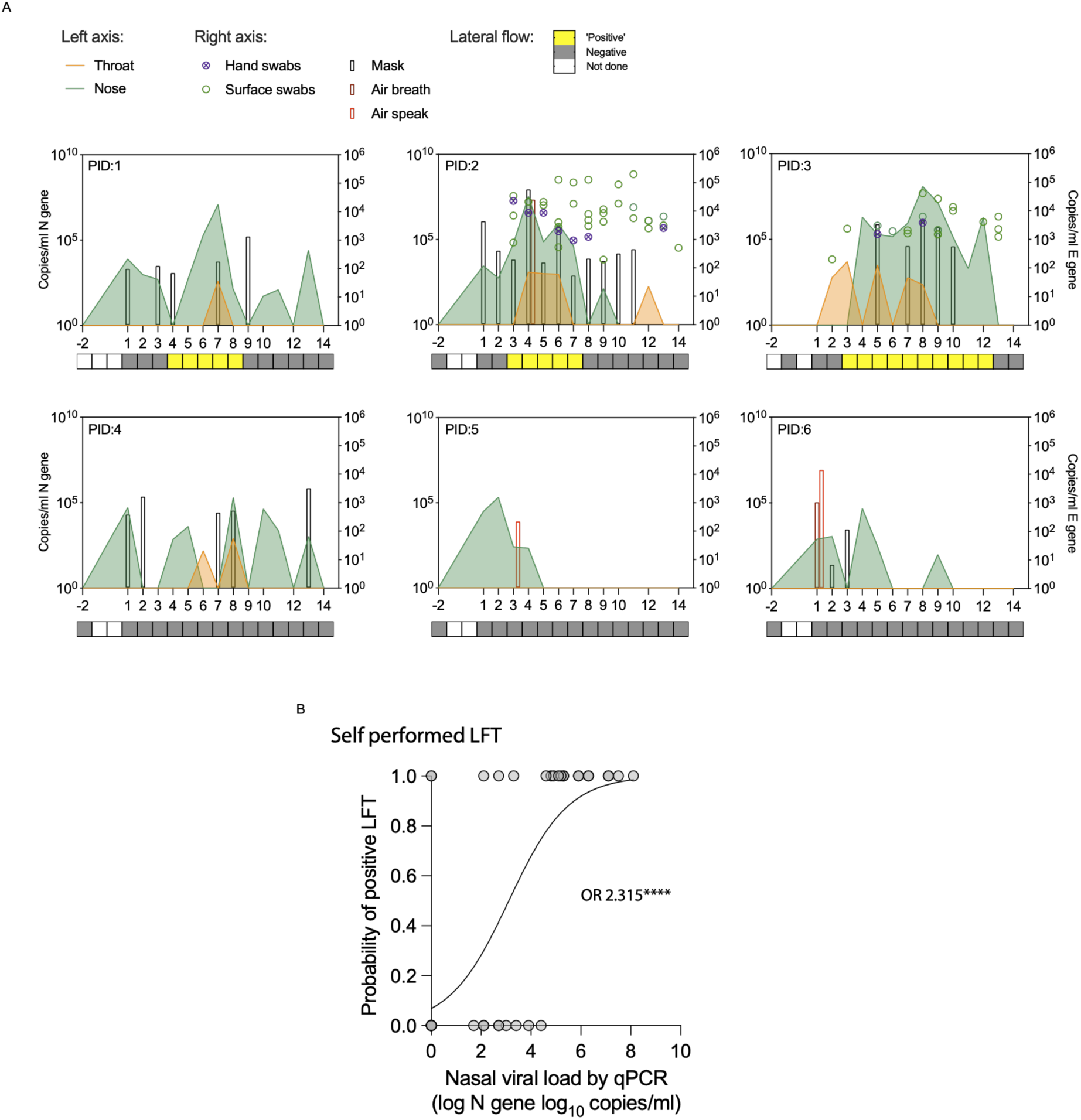
Highest frequency of emitted virus and positive lateral flow tests in symptomatic infected participants. **a**, Copies/ml SARS-CoV-2 N gene in swabs of the nose and throat (left axis) are overlayed with the copies/ml SARS-CoV-2 E gene from face masks, air samples and environmental swabs (right axis) measured by qPCR; 3 infected symptomatic (top row) and 3 infected asymptomatic (bottom row) participants. Results from daily participant self-performed lateral flow tests are shown underneath each participant graph. Logistic regression of **b**, self-performed LFT positive probability vs qPCR viral load. ****p=<0.0001.

Participants self-performed LFT with self-collected nasal swabs during the quarantine period. Overall, self-performed LFT positivity was associated with periods when URT VL was high and when virus was being emitted into the air and environment. Logistic regression modelling to assess the relationship between VL and LFT showed nasal VL as a significant predictor of LFT positivity (odds ratio 2.315, 95% CI 1.594-3.905; p=<0.0001) (Fig. 3b).

### Delta breakthrough infection consistently induces weak T cell responses

Following sustained infection, IFN-g producing T cells against overlapping nucleocapsid- (N), spike- (S) and membrane- (M) peptides were increased at day 7 p.i. compared with baseline by ELISpot. Median increases to 272.4 SFU/million PBMC against N, 567.2 SFU/million PBMC against S and 56.0 SFU/million PBMC against M (Fig. 4a, b, c) occurred, but only M reached significance (p=0.0215). No significant increases in T cells were detectable in the infected group when stimulated with separate MHC class I- (CD8) and class II- (CD4) restricted predicted epitope pools (Supplementary Fig. 2a, b). There was a trend towards a boost in T cell frequencies against N and M in the transient group, with some individuals showing a substantial increase between day 0 and 7, although this did not reach statistical significance (Supplementary Fig. 2c). No T cell responses were observed in the uninfected group and there was no difference in T cell response level between symptomatic vs asymptomatically infected participants. Thus, modest increases T cell responses, particularly non-S, were induced by mild virologically-confirmed infection with the Delta variant, with limited boosting by transient infection.

**Fig. 4.**
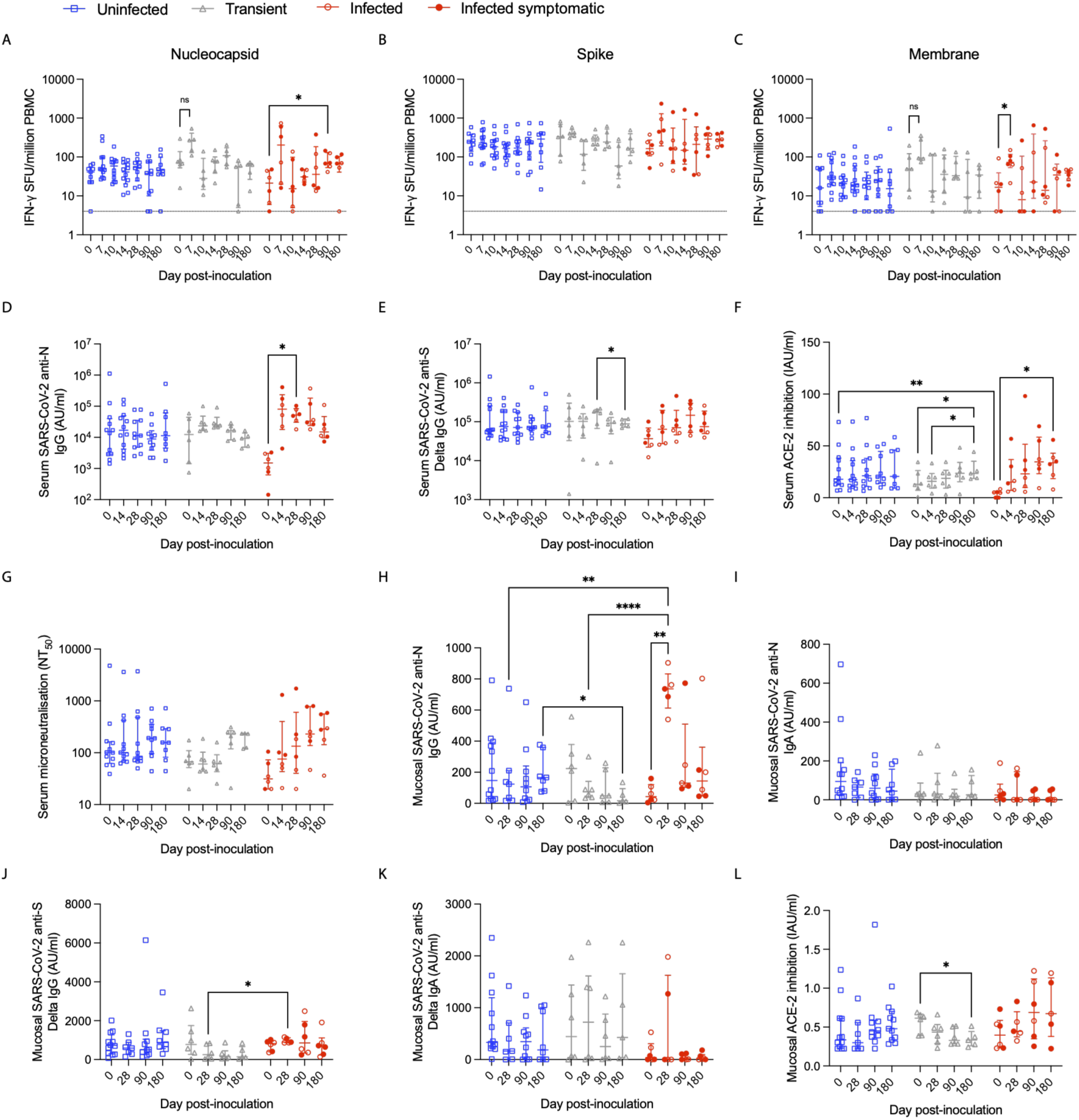
Weak T cell responses post-inoculation. T cell IFN-g spot forming units (SFU) per million PBMC in response to **a**, nucleocapsid, **b**, spike and **c**, membrane overlapping peptides pre and post-inoculation in the 3 infection outcome groups (uninfected n=12, transient n=6 and infected n=6). Line at lower limit of detection (LLOD). Serum **d**, anti-N IgG, **e**, anti-S IgG, **f**, ACE-2 inhibition and **g**, microneutralisation measured in the serum before and after inoculation. Mucosal **h**, anti-N IgG, **i**, anti-N IgA, **j**, anti-S IgG and **k**, anti-S IgA, and **l**, ACE-2 inhibition were measured in the nasal lining fluid. Three day 180 timepoints were excluded due to the participant contracting SARS-CoV-2 or OC43 in the community prior to this visit. Two-Way ANOVA mixed effects model with Geisser-Greenhouse correction and Tukey’s multiple comparisons test. Median and IQR. *p=<0.05, **p=<0.01, ****p=<0.0001.

### Boosting of anti-N IgG and neutralising antibody responses after breakthrough infection

Next, we measured antibody responses in serum and nasal lining fluid (NLF). In serum, there was a significant increase in anti-N IgG in the infected group at day 28 compared with baseline (median 52,072 AU/ml (IQR 30,623-84,021) at day 28 vs 1507 AU/ml (IQR 621-3050) at baseline, p=0.0374) (Fig. 4d). However, rises in serum anti-S IgG levels after infection were not significant (Fig. 4e). Both anti-N and anti-S serum IgG had begun to decline at day 90 (Fig. 4d, e). In two transiently infected individuals with the lowest anti-N IgG baseline antibody levels, a boost in serum anti-N IgG was also observed post-infection (Supplementary Fig. 2d). These two participants also had high fold-changes in anti-N T cell response between day 0 and 7. SARS-CoV-2 specific antibody was measured by microneutralisation assay (MNA) and ACE-2 inhibition (ACE-2i), the latter as a rapid-turnaround surrogate for virus neutralisation. In the infected and transient group, there were modest but significant rises in ACE-2i at day 180 compared with baseline, that was greater in the infected (median increases infected; 28.8IAU/ml, transient; 10.58 IAU/ml) (Fig. 4f). A non-significant trend toward increasing MNA antibody post-infection was observed in the infected group (Fig. 4g).

In NLF, anti-N IgG rose significantly in infected participants at day 28 with no similar increase in uninfected and transient groups (Fig. 4h). There was no increase in anti-N IgA (Fig. 4i). Modest rises in NLF anti-S IgG and IgA were seen in some infected participants p.i, and there were significantly higher anti-S IgG levels at day 28 in infected vs transient groups (Fig. 4j, k). ACE-2i levels were low in NLF with no increase p.i (Fig. 4l). No significant increases in NLF antibody were detected in transiently infected participants (Supplementary Fig. 2e). Therefore, despite the stringent virological criteria defining breakthrough infection, subsequent immune responses were overall of low magnitude and heterogeneous, consistent with the varying antigenic exposure histories of each individual and resulting immune backgrounds.

### Serum and mucosal antibody are associated with protection from infection

With the requirement for seroscreening to permit breakthrough infection already implying an association between serum neutralising antibodies and risk of infection, we sought to further pinpoint potential correlates of protection. 35% of all participants attending pre-screening had NT_50_ ≤1:80 and it had therefore been selected as a pragmatic threshold for seroselection. Applying more stringent NT_50_ cut-offs would have reduced numbers of suitable candidates (of all pre-screening volunteers, 19.3% had NT_50_ ≤1:40, 7.6% had NT_50_ ≤1:20). Pre-screening NT_50_, anti-S and anti-N binding antibody levels were significantly lower in infected participants than the overall pre-screened cohort (Fig. 5a). All participants who developed breakthrough infection had NT_50_ ≤1:40, with those who were symptomatic having undetectable NT_50_ (≤1:20) and undetectable anti-N IgG (≤1.00) (Table 1 and Fig. 5a). Transiently infected participants had an intermediate NT_50_ level (median=1:53 (IQR 1:87)) between those who became infected (median=1:20 (IQR 1:20)) and those who were uninfected (median=1:80 (IQR=1:310)).

**Fig. 5.**
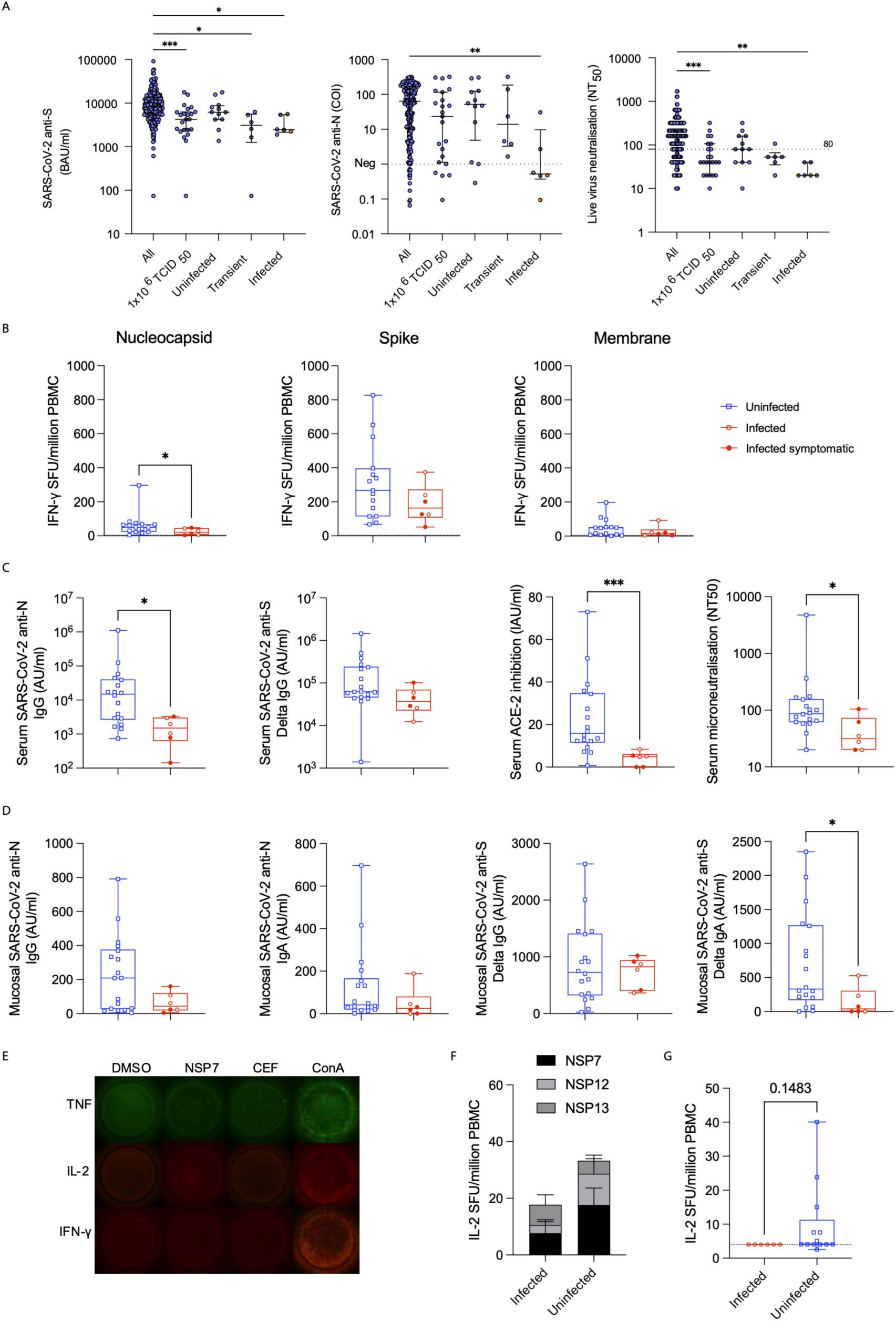
Serum and mucosal antibody are associated with protection from infection. **a**, Serum anti-spike (S) and anti-nucleocapsid (N) measured by Roche ECL assay and live virus neutralising antibody measured at the pre-screening visit split by study or outcome group (all pre-screened n=236, 1x10^6^ TCID^50^ group n=24, uninfected n=12, transient n=6 and infected n=6). Infected symptomatic participants are coloured in orange. Dotted line shows the negative cut off for N antibody level on the Roche ECL assay and 1:80 on the live virus neutralising antibody (seroscreening cut off). **b**, Baseline T cell IFN-g responses measured by ELISpot in response to N, S, M peptide pools (n=6 infected, n=6 transient and n=12 uninfected). Line at LLOD. **c**, Baseline serum anti-N IgG, anti-S IgG, ACE-2 inhibition, microneutralisation and **d**, mucosal anti-N IgG, anti-N IgA, anti-S IgG and anti-S IgA measured in the NLF (infected n=6 and uninfected n=18). **e**, Fluorospot representative images of baseline IL-2 producing T cells in response to **f**, NSP7, NSP12 and NSP13 (infected (n=6) and a combined uninfected group with the transient infected group included (n=15)) and **g**, CD45RE peptide pools (infected n=6 and uninfected n=14). Line at LLOD. One-Way ANOVA Kruskal-Wallis test with Dunn’s multiple comparisons test (a) or Mann-Whitney U test. Median and IQR. *p=<0.05, **p=<0.01, ***p=0.001.

Baseline immune measures were further compared between the infected and a combined uninfected group (including both transiently infected and uninfected, representative of individuals who would have been considered essentially uninfected in a community setting). This combined uninfected group had significantly higher N-specific T cell frequencies (Fig. 5b), serum anti-N IgG, ACE-2i and MNA antibody (Fig. 5c), and NLF anti-S IgA (Fig. 5d) at baseline compared with the infected group.

Previously we had shown a trend towards higher IL-2 producing T cells specific for proteins of the replication transcription complex (RTC) of SARS-CoV-2 (NSP7, 12, 13) in the group who resisted infection after pre-Alpha SARS-CoV-2 challenge of seronegative adults. We speculated that these T cells, likely primed by previous seasonal coronavirus infections, were able mount rapid responses, aborting infection^10^. In this study, NSP7/12/13-specific T cell IL- 2, TNF but not IFN-g responses measured by Fluorospot (representative images in Fig. 5e) were also enriched in the uninfected group compared with the infected group at baseline, but this did not reach significance (Fig. 5f and Supplementary Fig 3a, b). As T cell responses p.i. were limited to non-S antigens, a predicted epitope peptide pool of non-S peptides (CD4RE^11^) was also used for stimulation and again showed non-significant enrichment of IL-2 producing T cell responses in the uninfected group (Fig. 5g).

Comparative analysis was also performed using the three sub-groups; infected, transiently infected, and uninfected. Baseline N-specific T cell responses in the per protocol infection group were significantly lower than the transiently infected group, which itself had a higher median level than the uninfected group overall (Supplementary Fig. 3c). This suggests that N-specific T cell responses may play a role in protection from sustained infection but does not explain why transiently infected participants were not fully protected. Participants with transient infection had anti-N and anti-S antibody levels more akin to those uninfected, whereas serum ACE-2i and MNA antibody titres were intermediate between the infected and uninfected groups, suggesting a role for neutralising antibody as a stronger predictor of complete protection (Supplementary Fig. 3d, e).

### Baseline humoral immune measures strongly predict clinical outcomes after SARS-CoV-2 Delta challenge

Breakthrough infection occurs in the face of complex pre-existing immunity that differs between individuals, enhanced by varying degrees of previous vaccination and infection. Therefore, while serum antibodies were the strongest predictors of upper respiratory tract protection, they may actually represent surrogates for true mechanistic correlates of protection. To further investigate this, correlation analysis of all baseline immune features against each other was undertaken (Fig. 6a). Unsurprisingly, a number of serum antibody measures, including MNA, ACE-2i and anti-S binding antibodies, were co-correlated. In addition, serum ACE-2i and MNA antibody strongly correlated with nasal anti-S IgG and IgA, and anti-S IgA, while serum anti-S and anti-N IgG correlated with their respective nasal IgA antibodies. Serum anti-N IgG also correlated with the CD8 predicted epitope pool T cell response and several other antibody measurements correlated with T cell measurements, likely due to coordinated induction of these responses by previous infection.

**Fig. 6.**
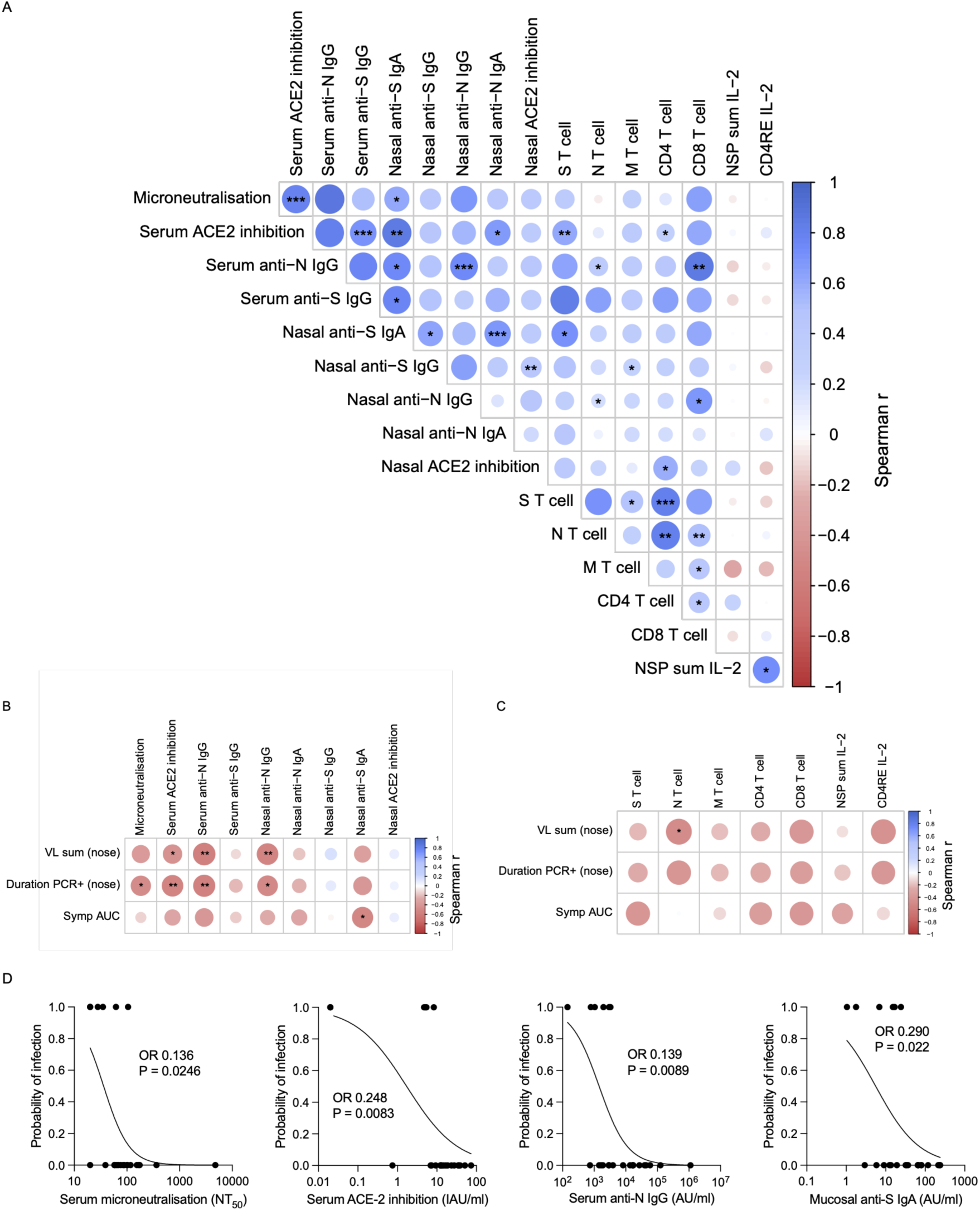
Baseline immunity correlates with outcome. **a**, Spearman correlation matrix between all baseline immune features and sum nasal log_10_ N gene copies/ml, symptom score AUC and duration qPCR positive (uncorrected for multiple comparisons). N=20 in total for correlation matrix (including all 6 infected). Correlation of baseline **b**, antibody and **c**, T cell immune features and infection outcomes. **d**, logistic regression plots for the 4 significant immunological markers that strongly predicted complete protection, odds ratios (OR) show standardised OR (see Table 2). *p=<0.05, **p=<0.01, ***p=<0.001.

**Table 2:** Univariate associations of baseline immunological predictors with infection outcome (with Firth’s penalised logistic regression to account for the small number of events). Odds ratios (ORs) and 95% confidence intervals (CIs) are standardised per one standard deviation increase in the predictor.

| Immune feature | Standardised OR | 95% CI | P value |
| --- | --- | --- | --- |
| Serum microneutralisation (MNA) | 0.136 | 0.013-0.837 | **0.0264** |
| Serum ACE-2 inhibition | 0.248 | 0.008-0.751 | **0.0083** |
| Serum anti-N IgG | 0.139 | 0.006-0.687 | **0.0089** |
| Mucosal anti-S IgA | 0.290 | 0.06-0.860 | **0.022** |
| Serum anti-S IgG | 0.570 | 0.168-1.382 | 0.217 |
| Mucosal anti-N IgA | 0.501 | 0.163-1.226 | 0.132 |
| Mucosal anti-S IgG | 1.103 | 0.475-3.375 | 0.827 |
| Mucosal anti-N IgG | 0.519 | 0.180-1.265 | 0.151 |
| Mucosal ACE-2 inhibition | 0.729 | 0.258-1.811 | 0.502 |
| S T cell | 0.642 | 0.231-1.566 | 0.334 |
| N T cell | 0.392 | 0.107-1.073 | 0.069 |
| M T cell | 0.606 | 0.178-1.77 | 0.365 |
| CD4 T cell | 0.559 | 0.194 0 1.347 | 0.198 |
| CD8 T cell | 0.51 | 0.155-1.483 | 0.218 |

To investigate the relationship between baseline immune features with VL and symptom outcomes, nasal N gene Log_10_ copies/ml were summed across all timepoints (including day 1 onwards), and VL duration (number of days positive for nasal N gene) and symptom score AUC were calculated (Supplementary Fig.1a). Correlation analysis of baseline measures thus revealed significant negative correlations between pre-inoculation immune features and VL or symptom scores (Fig. 6b, c). In particular, higher MNA antibody, serum anti-N IgG and nasal anti-N IgG levels at baseline were associated with lower VL and shorter duration of infection. Nasal anti-S IgA was associated with a lower symptom score AUC (Fig. 6b). Higher N specific T cell response at baseline was also associated with lower VL magnitude (Fig. 6c).

Univariate logistic regression analyses (with correction for small number of events) therefore identified 4 baseline immunological markers that strongly predicted complete protection (Table 2). Serum anti-N IgG levels (p=0.0089), MNA (p=0.026), serum ACE-2i (p=0.0083) and NLF anti-S IgA (p=0.022) all showed significant standardised odds ratios (Table 2 and logistic regression plots shown in Fig. 6d). While circulating T cell frequencies did not reach statistical significance, it remains a possibility across a larger population that these factors could also represent explanatory variables for protection from infection as well as disease.

## Discussion

While previous SARS-CoV-2 human challenge of seronegative volunteers readily resulted in symptomatic infection, this study for the first time models sustained breakthrough infection with mild-to-moderate Delta SARS-CoV-2 infection. Infection was safely induced in a third of healthy adults. High-dose inoculation and selection for low pre-existing serum neutralising antibody level was necessary. The low prevalence of serosusceptible individuals likely explains why earlier variants such as Delta were rapidly replaced in view of such intense immune pressure. Nevertheless, this study provides proof-of-principle that experimentally induced breakthrough infection can enable discovery of correlates of protection despite the high levels of pre-existing immunity across almost the entire population.

Following breakthrough infection, viral dynamics, viral emissions and symptom profiles were highly variable. This reflects the natural variation in infection outcomes, as well as the varied immune backgrounds that now exist between individuals due to vaccines, natural infections and differential boosting or waning of immune responses. Only 50% of infections were symptomatic, in line with data suggesting a high asymptomatic fraction since the introduction of SARS-CoV-2 vaccination^12^. Breakthrough Delta experimental infection also demonstrated substantially lower viral loads and shorter periods of infectivity than primary infection after pre-Alpha SARS-CoV-2 challenge of seronegative individuals^8^. Similarly, anosmia was no longer common, occurring in the single participant with the highest viral load peak and greatest duration of infectivity, in line with other studies^8^. Those who were symptomatic showed a trend towards being more infectious, shedding more virus in the URT, emitting more virus and with longer duration of LFT positivity. This is in contrast with data from the seronegative pre-Alpha challenge study suggesting symptom burden was not the main driver of infectiousness^6^.

In those who were infected or transiently infected, post-inoculation nucleocapsid antibody and T cell responses were more marked than spike responses, and there was minimal boosting of neutralising antibody. This contrasts with studies of T cell and antibody kinetics after Delta and Omicron breakthrough infection that have shown rapid recall of S-specific T cells as early as day 1 post-symptom onset and boosting of neutralising antibody^13^. We postulate that spike-specific responses had already reached near-maximum in this highly vaccine-experienced, young adult group not long after the peak of the pandemic. In contrast, the infected group had baseline serum anti-N IgG at or below levels determined to be negative. Anti-N antibodies have been shown to wane rapidly following natural infection^14^. Therefore, absence of previous infection or anti-N antibody waning since the last infection could explain negative nucleocapsid antibody levels, resulting in more headroom for anti-N responses. Indeed, we observed that infection-induced antibody responses began to wane or plateau from as early as day 28 in line with the kinetics of primary responses after pre-Alpha challenge of seronegative adults^5^.

Similar to the previous pre-Alpha SARS-CoV-2 challenge studies, transient infections occurred frequently, suggesting rapid control by host immunity in many individuals^4,7^. These transient infections are highly heterogenous, usually asymptomatic, and only induce weak, local or no immune response. Transient infection has been noted in community infections with a low rate of specific cellular or humoral immune responses but studies able to capture these are rare. For example, twice-weekly PCR testing of football league players revealed single positive PCRs in 11 of 98 positive individuals, they were all asymptomatic with only 1 mounting an immune response^15^. In the pre-Alpha study, transient infection of seronegative individuals was associated with a rapid (day 1) increase in nasopharyngeal innate and adaptive immune cells, absent in those who developed sustained infection, thus implying an immediate local protective immune response. In the pre-Alpha seropositive study^4^, transient infections were associated with lower baseline T cell frequencies to the same CD8 peptide pool used in this study and lower pre-existing serum and mucosal antibody levels compared with the uninfected outcome. This is supported by the findings here, where higher baseline N-specific T cell responses were also seen in transiently infected participants compared with sustained infection.

While serum neutralising antibody has been well established as a correlate of protection from disease, it does not fully explain protection from infection^1,16^. Here, humoral immune markers were more strongly predictive of protection from breakthrough infection than T cell responses, even after pre-selection of participants for low serum neutralising antibody titres. While this might reflect more immediate elimination of infectious virus by antibody, it is also well recognised that assays of cell-mediated and mucosal immunity are intrinsically more variable and difficult to standardise. Nevertheless, higher blood N-specific T cell responses and mucosal anti-N IgG level were associated with lower viral load and mucosal anti-S IgA was associated with lower symptom scores and protection. Mucosal IgA antibody has previously been shown to correlate with protection against upper respiratory infection in controlled human infection models of other respiratory viral infections^17^ and animal studies^18^, together highlighting the potential for boosting upper airway immunity to induce sterilising immunity and block transmission. Mucosal T cell responses are also implicated in early immune protection from infection and reducing viral load^7,10,19^. Furthermore, the consistent finding of anti-N responses associated with protection reaffirms that factors derived from infection (or hybrid immunity) may be responsible for the greater capacity to prevent breakthrough infection at population level^20^.

This study commenced before emergence of the Omicron variant. However, its findings demonstrate the feasibility of modelling breakthrough infection by SARS-CoV-2. Further development of controlled human infection models with Omicron variants will establish not only a platform for testing of next-generation interventions but also greater delineation of the immune factors critical for preventing infection itself and epidemic/pandemic spread.

## Data Availability

All data produced in the present study are available upon reasonable request to the authors

## Author Contributions

AS, HRW, HM, CC conceived or designed the study. AS, LS, PF, OD, MB, MSW, JML, AM, RLR, EH, HPJ, JZ, KS, MM, HM, CC conducted the clinical study or collected samples and data. AS, HRW, JZ, AB, SPS, MY, SAH, AA, BN, FS, DSK, JG, MG, SL, IS, KS, MM, AR, JM, HS, CE, SA, WB, RST, MC, MPD, GPT, HM, CC processed samples or acquired and analysed data. AS and HRW drafted the manuscript. CC critically revised the manuscript. All authors had full access to all the data in the study and had final responsibility for the decision to submit for publication.

## Data Sharing statement

De-identified participant data will be made available upon requests directed to the chief investigator. Requests should be made to the corresponding author. After approval of a proposal, data can be shared through a secure online platform after signing a data access agreement.

## Acknowledgements

We thank the study participants for their time and commitment. This study was funded by the Wellcome Trust. AS was supported by an NIHR Academic Clinical Lectureship. CC, AS and GPT were supported by the NIHR Imperial Biomedical Research Centre (BRC) award to Imperial College Healthcare NHS Trust and Imperial College. Infrastructure support was provided by the NIHR Imperial BRC and the NIHR Imperial Clinical Research Facility. We thank Ashley Otter and team at the UK Health Security Agency (UKHSA) for serological analysis of pre-screening serum for anti-S and anti-N IgG. MPD is supported by an NHMRC Investigator grant (#2034108).

