## Supplementary Data for "Biomarkers of protection against controlled human SARS-CoV-2 Delta variant breakthrough infection"

### Supplementary Material

**Supplementary Methods**

Study design

The study was conducted in high containment units at the Chelsea and Westminster NHS Foundation Trust and Oxford Experimental Medicine Clinical Research Facility (EMCRF). Participants were quarantined in single occupancy, negative pressure side rooms. Infected individuals remained in quarantine for a minimum of 14 days, while those who were uninfected stayed for 10 days, until discharge criteria were met (see study protocol for criteria). Participants were followed up for 1 year post-inoculation.

Procedures

Participants were inoculated intranasally by pipette. For dosing groups $10^2$-$10^5$ 50% tissue culture infectious dose ($TCID_{50}$), an inoculum volume of 200ul (4 drops of 50ul, alternating nostrils) was used. For dosing group $10^6$ $TCID_{50}$, inoculum volume of 516ul (6 drops of 86ul, alternating nostrils) was used.

The per protocol definition of infection was quantifiable RT-PCR detection of E or N gene greater than the lower limit of quantification (LLOQ) from throat or mid-turbinate swabs on 2 consecutive days after day 2. Transient infection (exploratory) was determined as any quantifiable RT-PCR detection greater than the LLOQ from throat or mid-turbinate swabs on non-consecutive days excluding day 1. Successive cohorts of 5-10 participants were intranasally inoculated with doses starting from $1x10^2$ $TCID_{50}$ to $1x10^6$ $TCID_{50}$, with dose escalation occurring if an attack rate of ≥50% was not achieved. Seroselection was implemented in the final $1x10^6$ $TCID_{50}$ cohort, for live virus neutralising antibody titre ($NT_{50}$) of ≤1:80 in serum. One volunteer in the $1x10^6$ $TCID_{50}$ group was lost to follow up after day 14.

Throat and mid-turbinate swabs were collected on day -2 pre-inoculation and then once daily from day 1 post-inoculation using flocked swabs. Swabs were placed into 3ml virus transport media (VTM) (BioServ, UK) and used for viral load measurements by RT-qPCR, plaque assay and antigen detection by lateral flow antigen test (LFT) (Flowflex™). LFT (Flowflex™) were also self-performed daily by participants themselves, following manufacturer's written instructions, at a pre-inoculation timepoint (day -2 ,-1 or day 0 pre-inoculation) and daily during the quarantine period.

Serum (serum separating tubes) was collected, isolated by centrifugation and frozen at pre-screening, baseline (day -1 -2 or 0 pre-inoculation; denoted day 0 on graphs), 14, 28, 90 and 180. Nasosorption samples were collected using synthetic absorptive matrix (SAM) strips (Nasosorption FX-I-11, Hunt Developments, Midhurst, UK) at day 0 and daily during the quarantine period, then at day 28, 90 and 180. SAM strips were snap frozen on dry ice and stored at -80°C, eluted into 330µl Millipore assay buffer AB-33k supplemented with 1% (v/v) Triton-X100 (to ensure SARS-CoV-2 inactivation) and aliquots frozen at -80°C before use.

Venous blood (lithium heparin) samples were collected at baseline (day -1, -2 or 0 pre-inoculation; denoted day 0 on graphs), day 7, 10, 14, 28, 90 and 180. PBMC were isolated using the Histopaque method; whole blood samples were diluted 1:1 in PBS and overlayered onto Histopaque and centrifuged at 400xg for 30 minutes. Or Leucosep method; blood samples were poured into the Leucosep tube containing Lymphoprep and centrifuged at 1000xg for 13 minutes. Isolated PBMCs were washed in PBS, counted using a Countess automated cell counter (Thermo Fisher Scientific) and used immediately for ELISpot or cryopreserved for Fluorospot.

Participants maintained a daily symptom diary during quarantine, recording and scoring the severity of lower respiratory, upper respiratory and systemic symptoms three times a day. Grade 0, no symptoms; grade 1, just noticeable; grade 2, clearly bothersome from time to time but does not interfere with me doing my normal daily activities; grade 3, quite bothersome most or all of the time, and it stops me participating in activities. The daily symptom score was calculated by summing the individual scores for each symptom per day and the total score by summing across all days.

Smell disturbance was measured at day -1 pre-inoculation, day 1, 4, 7, 10, 13 during quarantine and at day 28 using the University of Pennsylvania Smell Identification Test (UPSIT), a validated test used in previous SARS-CoV-2 human challenge studies[1]. UPSITs were continued at day 90 and 180 if a change from baseline was observed. A change in score from baseline of over 4 points was considered abnormal.

Room air sampling was performed using a Coriolis µ air sampler (Bertin Technologies, France), approximately 1 meter from the participant's head. Air samples were collected for 10 minutes with normal breathing and activity, followed by 10 minutes whilst participants read the Rainbow Passage 3 times. Hand and environmental surface swabs (overbed table, bed frame, bedside table, television remote control, bathroom door, toilet flush, and sink tap handles) were collected daily. All samples were subjected to virological analyses (ie, RT-qPCR and virus culture) and also quantified for human housekeeping gene 18S rRNA by PCR, as previously described[2].

Two uninfected participants in the $1x10^6$ $TCID_{50}$ group contracted SARS-CoV-2 in the community before their day 180 visit, these timepoints are excluded from longitudinal immunology data analysis. One uninfected participant was confirmed to have an OC43 (seasonal betacoronavirus) infection (detected by BioFire) before their day 180 visit, their antibody and T cell response to SARS-CoV-2 spike (S) and nucleocapsid (N) increased at least 2-fold therefore this timepoint was also excluded.

Challenge Virus

The SARS-CoV-2 Delta variant challenge virus strain was originally obtained in mid-2021 from a nose-throat swab from an otherwise healthy young adult with mild COVID-19 in the community. The virus was isolated by inoculation with the clinical sample of a qualified cGMP Vero Cell line. Seed Virus Stocks for each virus were then generated by a further passage on the same cGMP Vero Cell line. The Zayed Centre for Research (ZCR) GMP manufacturing facility of Great Ormond Street Hospital (GOSH) subsequently used the Seed Virus Stocks to manufacture the Delta variant Challenge Virus in accordance with cGMP and produce a Challenge Virus Master Virus Bank. Individual person inoculum vials were then produced in accordance with cGMP by GOSH by dilution of the cGMP MVB with cGMP sucrose diluent. The challenge viruses have undergone extensive quality testing performed as part of the GMP manufacturing release processes according to pre-determined specifications (including identity, infectivity and contaminant / adventitious agent tests). The challenge viruses were stored in a secure –80°C freezer (normal temperature range –60°C to –90°C).

Virus quantification

Viral RNA extraction was carried out from nose and throat swabs collected in 3ml VTM following inactivation by lysis buffer containing guanidinium thiocyanate, guanidinium chloride and/or sodium dodecyl sulphate (SDS) from the relevant RNA extraction kit (Maxwell HT Viral TNA Kit, Promega, or QIAamp Viral RNA mini kit, Qiagen). RT-qPCR reactions were setup using 5µl of RNA in a 20µl reaction, consisting of 5µl of 4× TaqMan Fast virus one step Master Mix (ThermoFisher), and 2µl each of E gene, N gene and RNAse P primer/probe mix and 4µl nuclease-free water. Ct values were converted to copies per milliliter (copies/ml) as previously described[3]. Swabs with detection of viral RNA in only one duplicate were classed as negative, timepoints with either E or N gene detected in both duplicates were classed as positive. The N gene LLOQ is 275 copies/ml, values below the LLOQ that were detected are shown, values below the LLOQ that were not detected are assigned a value of 1 for visualisation purposes.

Virus in mask, air and environmental samples were quantified by E gene RT-qPCR. Nose and throat swabs, mask, air and environmental samples were also subjected to viral plaque assay, as previously described[2]. The plaque assay LLOD was 5 PFU/ml.

Antibody measurements

*Live virus neutralisation*

For participant seroselection, serology was performed on frozen serum aliquots collected at the pre-screening visit in real-time. The neutralisation capacity of pre-screening sera against the Delta challenge strain was evaluated using a live virus neutralisation assay on Vero cells expressing ACE-2 and TMPRSS2. Sera were serially diluted in duplicate in DMEM, incubated for 1 hour at room temperature with 100 $TCID_{50}$/well of SARS-CoV-2, and then transferred to 96-well plates pre-seeded with Vero cells. The plates were incubated at 37°C with 5% $CO_2$ for 72 hours. After incubation, the cells were fixed with 1% Crystal Violet, and the plates were read to maximum serum dilutions that can neutralise 100 $TCID_{50}$ virus[4].

*Electrochemiluminescence assays*

Pre-screening sera were additionally tested for anti-S and Anti-N IgG by Roche Electrochemiluminescence (ECL) assay at a UK Health Security Agency (UKHSA) laboratory. Roche anti-S was reported as binding antibody unit (BAU)/ml. Roche anti-N IgG was reported as cut-off index (COI) and values ≥1.00 are considered positive.

*Microneutralisation*

Baseline (day -2, -1 or day 0 pre-inoculation) and post-inoculation timepoint sera were tested by microneutralisation assay (MNA) performed as previously described[5]. Sera were serially diluted in DMEM supplemented with 1% fetal bovine serum (FBS) from an initial dilution of 1:10 to 1:10,000. Equal volumes of diluted sera and SARS-CoV-2 Delta virus (approximately 100 focus forming units) were combined and incubated for 30 minutes. Delta (B.1.617.2) isolate 83DJ-1 was provided by Piet Maes, Laboratory of Clinical and Epidemiological Virology (Rega Institute, Belgium). Following incubation, 100µl Vero E6 cells (supplied by UKHSA) ($4.5x10^5$/ml) was added to each well and virus was allowed to infect the cells for 2 hours at 37°C, 5% $CO_2$ followed by the addition of 100µl carboxymethyl cellulose (1%) to each well. The plates were incubated for a further 18-24 hours at 37°C, 5% $CO_2$. All assays were carried out in duplicate.

Cells were washed with 200µl DPBS and then fixed with paraformaldehyde 4% v/v (100µl/well) for 30 minutes at room temperature. Cells were permeabilised with Triton X100 (1% in PBS) and then stained for SARS-CoV-2 nucleoprotein using a human monoclonal antibody, FB9B[6].

Bound antibody was detected following incubation with a goat anti-human IgG HRP conjugate (Sigma, UK) and following TrueBlue Peroxidase substrate (Insight Biotechnology, London, UK) addition imaged using an ELISpot reader. The half-maximal inhibitory concentration ($NT_{50}$) was defined as the concentration of sera that reduced the foci forming unit (FFU) by 50% compared to the control wells.

*Meso Scale Discovery*

Serum and nasal lining fluid (NLF) IgG and IgA assays were performed using the Meso Scale Discovery (MSD) V-PLEX SARS-CoV-2 Key Variant Spike Panel 1 (IgG or IgA) Kits (Meso Scale Diagnostics, Rockville, USA). Reported antigens from these plates are Delta (B.1.617.2; AY.4 Alt Seq 2) SARS-CoV-2 S "SARS-CoV-2 anti-S Delta" and Nucleocapsid "SARS-CoV-2 anti-N".

The assays were performed according to the manufacturer's instructions, with all steps at room temperature and shaking incubations at 700 RPM. Briefly, the plates were blocked with Blocker A solution (30 minutes), followed by a wash step (three washes with 1X Wash Buffer), and a two-hour incubation with samples (diluted 1:1,000-10,000 in Diluent 100), Reference standard 1 (calibrator), and an internal assay control (Serology Control 1.1). The plates were washed, and the SULFO-TAG anti-human IgG/A antibody was added for 1 hour. Following this, the plates were washed and MSD GOLD Read Buffer B was added. The assays were read with the MESO QuickPlex SQ 120MM (model 1300) instrument, and the data analysed using the MSD Discovery Workbench software v4, with standard curves for each antigen created by fitting the signals from the reference standard using a 4-parameter logistic model. The IgG/A concentrations, expressed in Arbitrary Units (AU)/ml were determined from the electrochemiluminescence (ECL) signals by back-fitting to the standard curve and multiplying by the dilution factor.

Serum and NLF was also assessed for the ACE-2 inhibition ability (ACE-2i) of SARS-CoV-2 antibodies using the MSD pseudo-neutralisation assay with the V-PLEX SARS-CoV-2 Key Variant Spike Panel 1 (ACE2) Kit. The assays were performed according to the manufacturer's instructions. Briefly, wells were blocked with Blocker A solution (30 minutes), followed by a wash step and the addition of samples (diluted 1:10-1:100) and the ACE-2 Calibration Reagent 3 (8-point, 4-fold serial dilution standard starting with a 10-fold dilution, Cat. No. C01APE-2, A0080375) for 1 hour. Following this, SULFO-TAG Human ACE-2 Protein was added for an additional hour incubation. After washing, MSD GOLD Read Buffer B was added, and the assays were read with the MESO QuickPlex SQ 120MM (model 1300) instrument. The raw data was analysed with MSD Discovery Workbench software v4, and standard curves were created by fitting the signals from the standard (calibrator) using a 4-parameter logistic model. ACE-2 inhibition was presented as IAU/ml.

Cellular measurements

*ELISpot assay*

In brief, MultiScreen-IP ELISpot plates (Millipore) were coated with 1-D1K capture antibody (Mabtech) at room temperature for 3-8 hours, blocked with RPMI supplemented with 10% FBS at room temperature for 1-8 hours then $2.5x10^5$ PBMC per well in triplicate were plated per condition with peptides at 4ug/ml. Peptides were custom made Delta variant (challenge strain sequence) 15 mers overlapping by 11 of S subunit 1 and 2, membrane (M) and N proteins (Supplementary Table 2) (Think Peptides; Proimmune). Also tested were predicted SARS-CoV-2 $CD4^+$ and $CD8^+$ T-cell epitopes (La Jolla Institute for Immunology, San Diego, CA, USA[7]) (Supplementary Table 2). Plates were incubated at 37°C with 5% $CO_2$ for 18-20 hours. Anti-IFN-γ 7-B6-1-Biotin followed by SA-ALP and BCIP development was used for spot visualisation and numeration using an AID vSpot Spectrum Analyser. The mean of unstimulated (media) control wells was subtracted from the mean of peptide wells and spots were adjusted to spot forming units (SFU) per million PBMC. The lower limit of detection (LLOD) was 4 SPF/million PBMC.

*Fluorospot assay*

CTL (ImmunoSpot) fluorospot kit was run following the manufacturer instructions on a subset of samples indicated in the figure legends due to sample availability. Peptides covering the NSP7, NSP12 and NSP13 (Supplementary Table 3) at 1ug/ml and predicted 'remainder' (everything other than spike) pool CD4RE (La Jolla Institute for Immunology, San Diego, CA, USA[7]) were used. Internal plate controls were 2 DMSO wells (negative controls), concanavalin A (ConA; Sigma-Aldrich) and FEC (HLA I-restricted peptides from influenza, Epstein–Barr virus and CMV; 1 µg/ml per peptide; Miltenyi Biotec). Results were excluded where positive control wells (ConA and FEC) were negative. The average of the 2 DMSO wells was subtracted from the peptide stimulated wells for each sample and results were expressed as SFU/million PBMC. Any response lower in magnitude that 2s.d. of the average DMSO count was excluded. The lower limit of detection (LLOD) was 4 SPF/million PBMC.

Statistical analysis

To account for the small number of events even in the univariate analysis, Firth's penalised logistic regression was used. Prior to logistic regression, each immunological marker was standardised to have equal means and standard deviations (0 and 1 respectively on a $\log_{10}$ scale) to allow direct comparison of effect sizes across assays and markers. Standardisation was carried out using a left censored normal distribution to account for the observations below the limit of detection. Standardised odds ratios and 95% confidence intervals were reported, representing the expected relative change in odds of infection for every one-unit standard deviation increase in the amount of the immunological marker. All analyses were performed in *R (v.4.3.1)* using *logistf* package.

### Supplementary Figures

A

| Group | Participant | Pre-screeening live virus neutralisation (NT50) | Sum N gene Log10 copies/ml (nose) | Duration PCR positive (nose) (days) | Symptom score AUC |
|---|---|---|---|---|---|
| Infected | 1 | 20 | 35.0 | 10.0 | 108 |
| Infected | 2 | 20 | 33.9 | 7.0 | 94 |
| Infected | 3 | 20 | 52.6 | 9.0 | 51 |
| Infected | 4 | 20 | 27.5 | 7.0 | 2 |
| Infected | 5 | 40 | 9.8 | 2.0 | 1 |
| Infected | 6 | 40 | 13.1 | 4.0 | 0 |
| Transient | 7 | 107 | 8.5 | 2.0 | 6 |
| Transient | 8 | 40 | 5.7 | 2.0 | 17 |
| Transient | 9 | 40 | 0.0 | 0.0 | 29 |
| Transient | 10 | 53 | 2.7 | 1.0 | 53 |
| Transient | 11 | 53 | 6.4 | 2.0 | 9 |
| Transient | 12 | 20 | 1.8 | 1.0 | 53 |
| Uninfected | 13 | 213 | 4.0 | 1.0 | 12 |
| Uninfected | 14 | 10 | 2.6 | 1.0 | 2 |
| Uninfected | 15 | 107 | 0.0 | 0.0 | 12 |
| Uninfected | 16 | 40 | 0.0 | 0.0 | 3 |
| Uninfected | 17 | 160 | 0.0 | 0.0 | 0 |
| Uninfected | 18 | 40 | 0.0 | 0.0 | 32 |
| Uninfected | 19 | 40 | 1.9 | 1.0 | 0 |
| Uninfected | 20 | 80 | 0.0 | 0.0 | 21 |
| Uninfected | 21 | 80 | 3.3 | 1.0 | 12 |
| Uninfected | 22 | 320 | 0.0 | 0.0 | 0 |
| Uninfected | 23 | 160 | 4.5 | 1.0 | 35.5 |
| Uninfected | 24 | 80 | 2.5 | 1.0 | 4 |

N gene $Log_{10}$ copies/ml — Nose, Day post-inoculation

| Participant | -2 | 1 | 2 | 3 | 4 | 5 | 6 | 7 | 8 | 9 | 10 | 11 | 12 | 13 | 14 |
|---|---|---|---|---|---|---|---|---|---|---|---|---|---|---|---|
| 1 | | 3.9 | 3.0 | 2.7 | | 2.7 | **5.3** | **7.1** | 2.1 | | 1.7 | 2.1 | | 4.4 | |
| 2 | | 3.4 | 2.7 | **4.8** | **7.5** | 4.9 | 5.9 | 4.6 | | 2.1 | | | | | |
| 3 | | | | | **6.3** | **5.3** | **5.2** | **5.9** | **8.1** | **7.1** | 5.1 | 3.3 | 6.3 | | |
| 4 | | 4.7 | | | 2.9 | 3.6 | | | 5.3 | | 4.6 | 3.4 | | 3.0 | |
| 5 | | 4.5 | **5.3** | 2.4 | 2.3 | | | | | | | | | | |
| 6 | | 2.9 | **3.0** | | 4.7 | 2.5 | | | | 2.0 | | | | | |
| 7 | | 4.2 | 4.3 | | | | | | | | | | | | |
| 8 | | | 2.9 | | | | 2.8 | | | | | | | | |
| 9 | | | | | | | | | | | | | | | |
| 10 | | 2.7 | | | | | | | | | | | | | |
| 11 | | 3.8 | 2.7 | | | | | | | | | | | | |
| 12 | | 1.8 | | | | | | | | | | | | | |
| 13 | | 4.0 | | | | | | | | | | | | | |
| 14 | | 2.6 | | | | | | | | | | | | | |
| 15 | | | | | | | | | | | | | | | |
| 16 | | | | | | | | | | | | | | | |
| 17 | | | | | | | | | | | | | | | |
| 18 | | | | | | | | | | | | | | | |
| 19 | | 1.9 | | | | | | | | | | | | | |
| 20 | | | | | | | | | | | | | | | |
| 21 | | 3.3 | | | | | | | | | | | | | |
| 22 | | | | | | | | | | | | | | | |
| 23 | | 4.5 | | | | | | | | | | | | | |
| 24 | | 2.5 | | | | | | | | | | | | | |

N gene $Log_{10}$ copies/ml — Throat, Day post-inoculation

| Participant | -2 | 1 | 2 | 3 | 4 | 5 | 6 | 7 | 8 | 9 | 10 | 11 | 12 | 13 | 14 |
|---|---|---|---|---|---|---|---|---|---|---|---|---|---|---|---|
| 1 | | | | | | | | 2.6 | | | | | | | |
| 2 | | | | | **3.1** | 3.0 | 3.0 | | | | | | 2.2 | | |
| 3 | | | 2.8 | **3.7** | | **3.5** | | 2.8 | 2.4 | | | | | | |
| 4 | | | | | | | 2.2 | | 2.9 | | | | | | |
| 5 | | | | | | | | | | | | | | | |
| 6 | | | | | | | | | | | | | | | |
| 7 | | | | | | | | | | | | | | | |
| 8 | | **4.8** | | | | | | | | | | | | | |
| 9 | | | 3.4 | | | | | | | | | | | | |
| 10 | | | | 2.1 | | | | | | | | | | | |
| 11 | | | | | | | | | | | | | | | |
| 12 | | | 3.2 | 1.4 | | 1.4 | | | | | | | | | |
| 13 | | | | | | | | | | | | | | | |
| 14 | | | | | | | | | | | | | | | |
| 15 | | | | | | | | | | | | | | | |
| 16 | | | | | | | | | | | | | | | |
| 17 | | | | | | | | | | | | | | | |
| 18 | | 2.3 | | | | | | | | | | | | | |
| 19 | | 2.1 | | | | | | | | | | | | | |
| 20 | | | | | | | | | | | | | | | |
| 21 | | | | | | | | | | | | | | | |
| 22 | | | | | | | | | | | | | | | |
| 23 | | | | | | | | | | | | | | | |
| 24 | | | | | | | | | | | | | | | |

Key: Bold — Plaque positive; Negative; LLOQ; Not done

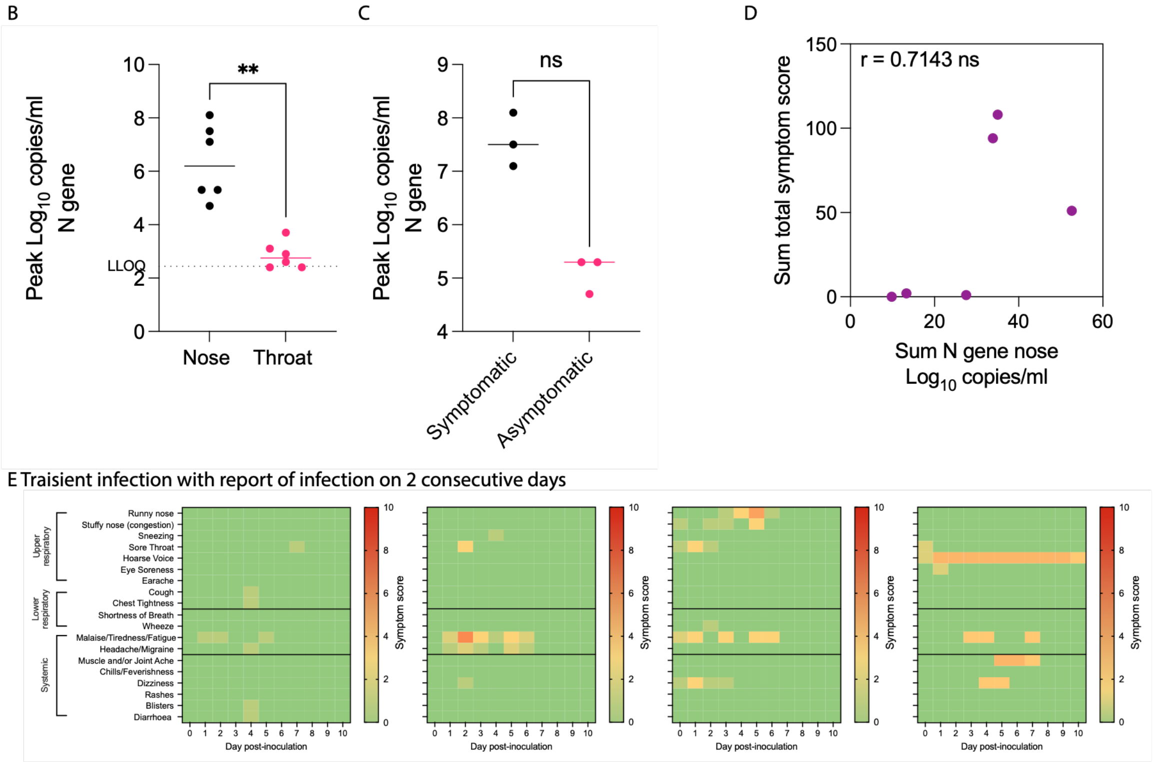

**Sup. Fig. 1. Viral load and symptom score comparisons.**

a, Heat map of the qPCR copies/ml SARS-CoV-2 N gene showing pre-screening live virus neutralisation titre and values used in correlation analysis in Fig. 6. b, Log10 copies/ml SARS-CoV-2 N gene in the nose vs the throat and c, in the symptomatic vs asymptomatic infected participants. c, Spearman correlation between the sum total symptom score and log10 copies/ml SARS-CoV-2 N gene in the nose in the infected group. e, Symptoms experienced by the transient group. Mann-Whitney U test. **p=<0.01.

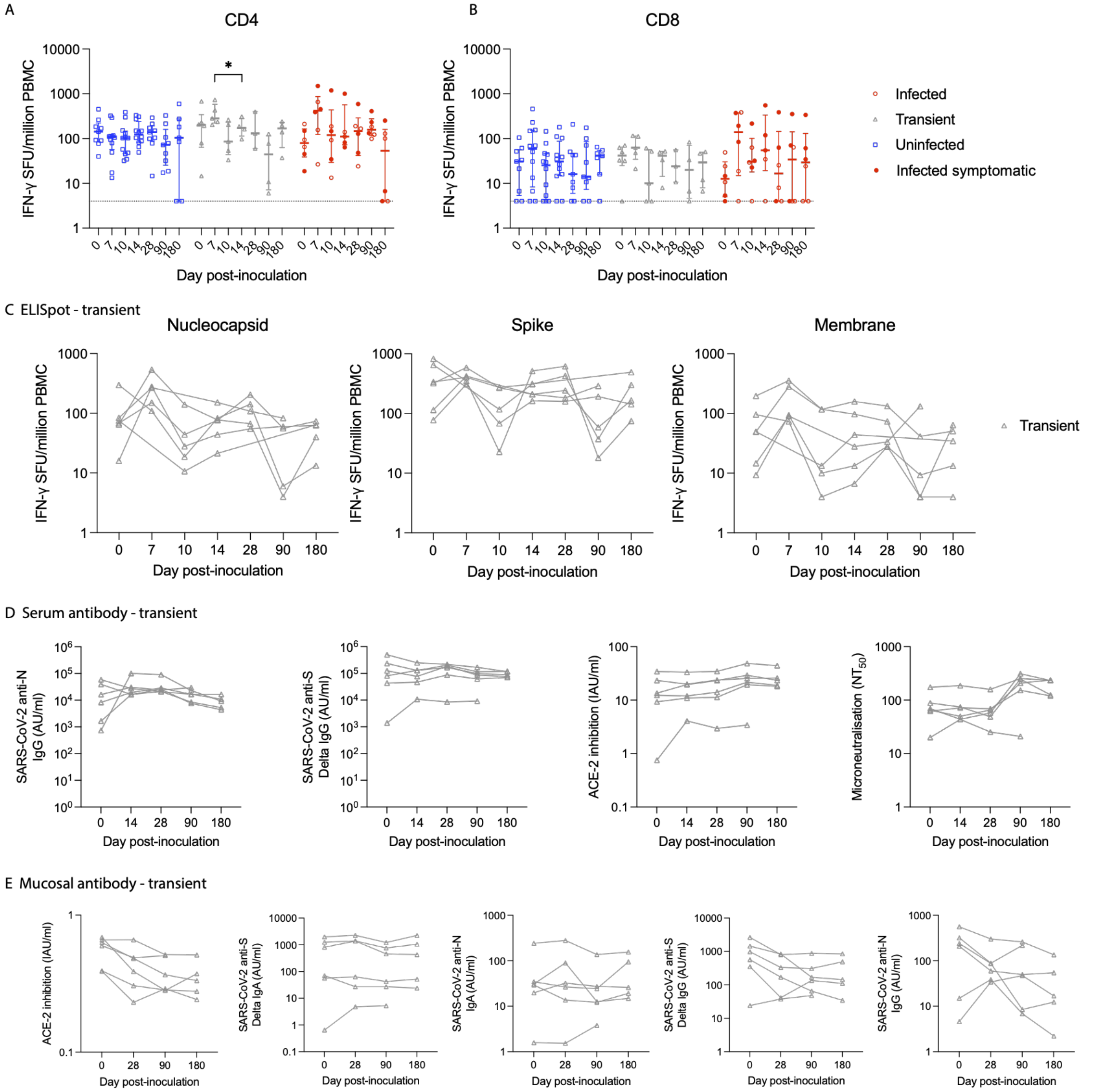

**Sup. Fig. 2. ELISpot and antibody responses in the transient group.**

T cell IFN-g spot forming units (SFU) per million PBMC in response to a, CD4 and b, CD8 predicted epitope peptide pool pre and post-inoculation in the 3 infection outcome groups (uninfected n=12, transient n=6 and infected n=6). c, T cell IFN-g spot forming units (SFU) per million PBMC in response to nucleocapsid, spike and membrane overlapping peptides pre and post-inoculation in the transient group (n=6). d, Serum anti-N IgG, anti-S IgG, ACE-2 inhibition and microneutralisation measured in the serum before and after inoculation in the transient group (n=6). e, Mucosal ACE-2 inhibition, anti-S IgA, anti-N IgA, anti-S IgG and anti-N IgG in the nasal lining fluid in the transient group (n=6). Three day 180 timepoints were excluded due to the participant contracting SARS-CoV-2 or OC43 in the community prior to this visit. Two-Way ANOVA mixed effects model with Geisser-Greenhouse correction and Tukey’s multiple comparisons test (a, b) or One-Way ANOVA Kruskal-Wallis test with Dunn’s multiple comparisons test (c-e). Median and IQR. *p=<0.05.

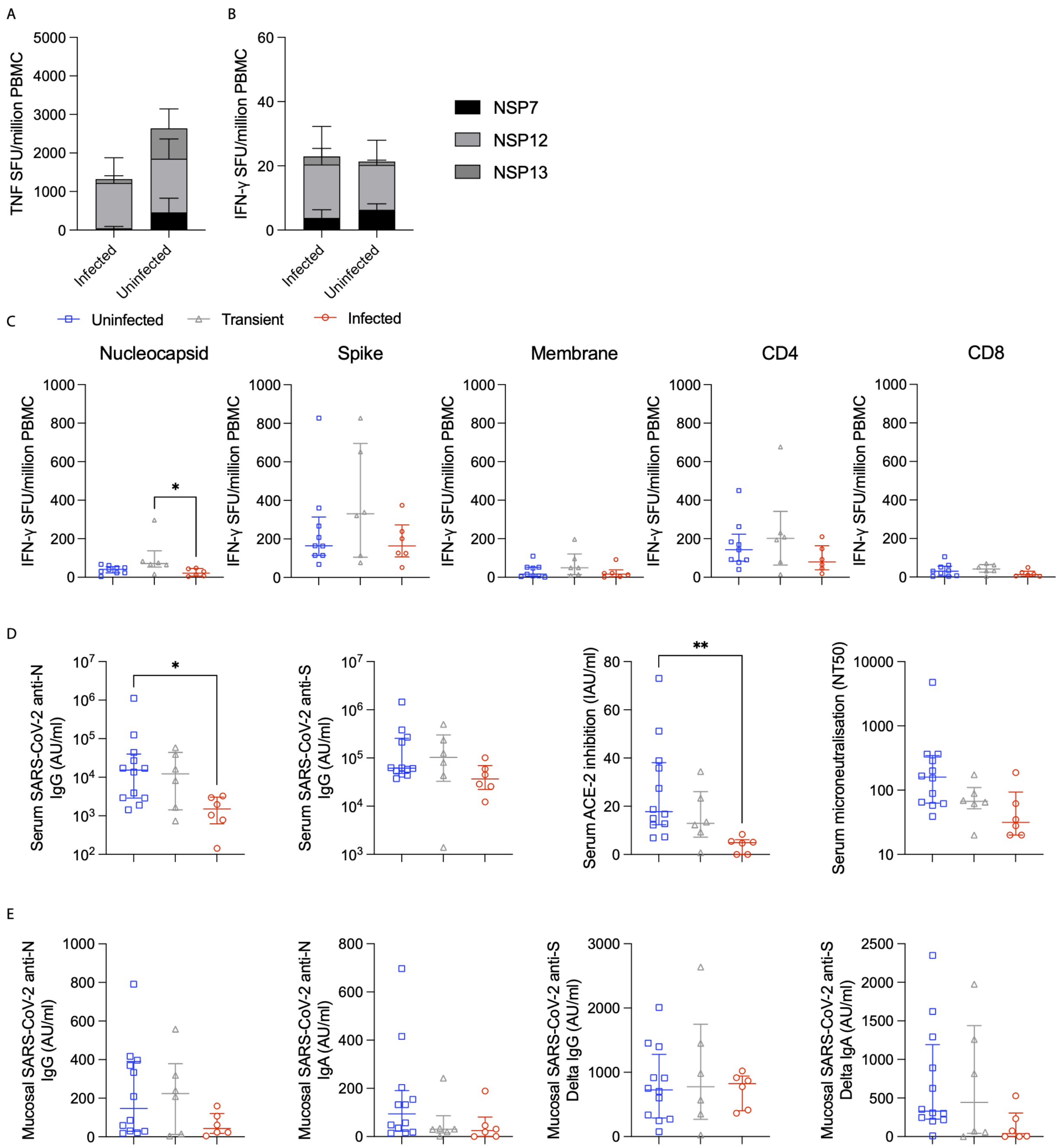

**Sup. Fig. 3. Serum and mucosal antibody are associated with protection from infection.**
a, TNF and b, IFN-g producing T cells at baseline in response to NSP7, NSP12 and NSP13 peptide pool stimulation split in to infected (n=6) and a combined uninfected group with the transient infected group included (n=15). c, Baseline T cell IFN-g responses measured by ELISpot in response to nucleocapsid, spike and membrane peptide pools split into the 3 groups (n=6 infected, n=6 transient and n=12 uninfected). d, Baseline serum anti-N IgG, anti-S IgG, ACE-2 inhibition, microneutralisation and e, mucosal anti-N IgG, anti-N IgA, anti-S IgG and anti-S IgA measured in the NLF. One-Way ANOVA Kruskal-Wallis test with Dunn's multiple comparisons test. Median and IQR. *p=<0.05, **p=<0.01.

**Supplementary Tables**

**Supplementary Table 1:** Adverse event deemed possibly related to challenge virus infection.

| Adverse event Description | Severity | Outcome | Serious Adverse Event (SAE) |
|---|---|---|---|
| Cold | Mild | Resolved | No |
| Feeling of mild tightness in throat | Mild | Resolved | No |
| Grade 1 systolic hypertension | Mild | Resolved | No |
| Grade 1 raised respiratory rate | Mild | Resolved | No |
| Malaise | Mild | Resolved | No |
| Headache | Mild | Resolved | No |

**Supplementary Table 2:** Structural antigen peptides used for *Ex vivo* IFN-γ T cell ELISpot assay.

| | Spike | | Nucleocapsid | | Membrane |
|---|---|---|---|---|---|
| 1 | MFVFLVLLPLVSSQC | 1 | MSDNGPQNQRNAPRI | 1 | MADSNGTITVEELKK |
| 2 | LVLLPLVSSQCVNLR | 2 | GPQNQRNAPRITFGG | 2 | NGTITVEELKKLLEQ |
| 3 | PLVSSQCVNLRTRTQ | 3 | QRNAPRITFGGPSDS | 3 | TVEELKKLLEQWNLV |
| 4 | SQCVNLRTRTQLPPA | 4 | PRITFGGPSDSTGSN | 4 | LKKLLEQWNLVIGFL |
| 5 | NLRTRTQLPPAYTNS | 5 | FGGPSDSTGSNQNGE | 5 | LEQWNLVIGFLFLTW |
| 6 | RTQLPPAYTNSFTRG | 6 | SDSTGSNQNGERSGA | 6 | NLVIGFLFLTWICLL |
| 7 | PPAYTNSFTRGVYYP | 7 | GSNQNGERSGARSKQ | 7 | GFLFLTWICLLQFAY |
| 8 | TNSFTRGVYYPDKVF | 8 | NGERSGARSKQRRPQ | 8 | LTWICLLQFAYANRN |
| 9 | TRGVYYPDKVFRSSV | 9 | SGARSKQRRPQGLPN | 9 | CLLQFAYANRNRFLY |
| 10 | YYPDKVFRSSVLHST | 10 | SKQRRPQGLPNNTAS | 10 | FAYANRNRFLYIIKL |
| 11 | KVFRSSVLHSTQDLF | 11 | RPQGLPNNTASWFTA | 11 | NRNRFLYIIKLIFLW |
| 12 | SSVLHSTQDLFLPFF | 12 | LPNNTASWFTALTQH | 12 | FLYIIKLIFLWLLWP |
| 13 | HSTQDLFLPFFSNVT | 13 | TASWFTALTQHGKEG | 13 | IKLIFLWLLWPVTLA |
| 14 | DLFLPFFSNVTWFHA | 14 | FTALTQHGKEGLKFP | 14 | FLWLLWPVTLACFVL |
| 15 | PFFSNVTWFHAIHVS | 15 | TQHGKEGLKFPRGQG | 15 | LWPVTLACFVLAAVY |
| 16 | NVTWFHAIHVSGTNG | 16 | KEGLKFPRGQGVPIN | 16 | TLACFVLAAVYRINW |
| 17 | FHAIHVSGTNGTKRF | 17 | KFPRGQGVPINTNSS | 17 | FVLAAVYRINWITGG |
| 18 | HVSGTNGTKRFDNPV | 18 | GQGVPINTNSSPDDQ | 18 | AVYRINWITGGIATA |
| 19 | TNGTKRFDNPVLPFN | 19 | PINTNSSPDDQIGYY | 19 | INWITGGIATAMACL |
| 20 | KRFDNPVLPFNDGVY | 20 | NSSPDDQIGYYRRAT | 20 | TGGIATAMACLVGLM |
| 21 | NPVLPFNDGVYFASI | 21 | DDQIGYYRRATRRIR | 21 | ATAMACLVGLMWLSY |
| 22 | PFNDGVYFASIEKSN | 22 | GYYRRATRRIRGGDG | 22 | ACLVGLMWLSYFIAS |

| | | | | | |
|---|---|---|---|---|---|
| 23 | GVYFASIEKSNIIRG | 23 | RATRRIRGGDGKMKD | 23 | GLMWLSYFIASFRLF |
| 24 | ASIEKSNIIRGWIFG | 24 | RIRGGDGKMKDLSPR | 24 | LSYFIASFRLFARTR |
| 25 | KSNIIRGWIFGTTLD | 25 | GDGKMKDLSPRWYFY | 25 | IASFRLFARTRSMWS |
| 26 | IRGWIFGTTLDSKTQ | 26 | MKDLSPRWYFYYLGT | 26 | RLFARTRSMWSFNPE |
| 27 | IFGTTLDSKTQSLLI | 27 | SPRWYFYYLGTGPEA | 27 | RTRSMWSFNPETNIL |
| 28 | TLDSKTQSLLIVNNA | 28 | YFYYLGTGPEAGLPY | 28 | MWSFNPETNILLNVP |
| 29 | KTQSLLIVNNATNVV | 29 | LGTGPEAGLPYGANK | 29 | NPETNILLNVPLHGT |
| 30 | LLIVNNATNVVIKVC | 30 | PEAGLPYGANKDGII | 30 | NILLNVPLHGTILTR |
| 31 | NNATNVVIKVCEFQF | 31 | LPYGANKDGIIWVAT | 31 | NVPLHGTILTRPLLE |
| 32 | NVVIKVCEFQFCNDP | 32 | ANKDGIIWVATEGAL | 32 | HGTILTRPLLESELV |
| 33 | KVCEFQFCNDPFLDV | 33 | GIIWVATEGALNTPK | 33 | LTRPLLESELVIGAV |
| 34 | FQFCNDPFLDVYYHK | 34 | VATEGALNTPKDHIG | 34 | LLESELVIGAVILRG |
| 35 | NDPFLDVYYHKNNKS | 35 | GALNTPKDHIGTRNP | 35 | ELVIGAVILRGHLRI |
| 36 | LDVYYHKNNKSWMES | 36 | TPKDHIGTRNPANNA | 36 | GAVILRGHLRIAGHH |
| 37 | YHKNNKSWMESGVYS | 37 | HIGTRNPANNAAIVL | 37 | LRGHLRIAGHHLGRC |
| 38 | NKSWMESGVYSSANN | 38 | RNPANNAAIVLQLPQ | 38 | LRIAGHHLGRCDIKD |
| 39 | MESGVYSSANNCTFE | 39 | NNAAIVLQLPQGTTL | 39 | GHHLGRCDIKDLPKE |
| 40 | VYSSANNCTFEYVSQ | 40 | IVLQLPQGTTLPKGF | 40 | GRCDIKDLPKEITVA |
| 41 | ANNCTFEYVSQPFLM | 41 | LPQGTTLPKGFYAEG | 41 | IKDLPKEITVATSRT |
| 42 | TFEYVSQPFLMDLEG | 42 | TTLPKGFYAEGSRGG | 42 | PKEITVATSRTLSYY |
| 43 | VSQPFLMDLEGKQGN | 43 | KGFYAEGSRGGSQAS | 43 | TVATSRTLSYYKLGA |
| 44 | FLMDLEGKQGNFKNL | 44 | AEGSRGGSQASSRSS | 44 | SRTLSYYKLGASQRV |
| 45 | LEGKQGNFKNLREFV | 45 | RGGSQASSRSSSRSR | 45 | SYYKLGASQRVAGDS |
| 46 | QGNFKNLREFVFKNI | 46 | QASSRSSSRSRNSSR | 46 | LGASQRVAGDSGFAA |
| 47 | KNLREFVFKNIDGYF | 47 | RSSSRSRNSSRNSTP | 47 | QRVAGDSGFAAYSRY |
| 48 | EFVFKNIDGYFKIYS | 48 | RSRNSSRNSTPGSSM | 48 | GDSGFAAYSRYRIGN |
| 49 | KNIDGYFKIYSKHTP | 49 | SSRNSTPGSSMGTSP | 49 | FAAYSRYRIGNYKLN |
| 50 | GYFKIYSKHTPINLV | 50 | STPGSSMGTSPARMA | 50 | SRYRIGNYKLNTDHS |
| 51 | IYSKHTPINLVRDLP | 51 | SSMGTSPARMAGNGC | 51 | IGNYKLNTDHSSSSD |
| 52 | HTPINLVRDLPQGFS | 52 | TSPARMAGNGCDAAL | 52 | KLNTDHSSSSDNIAL |
| 53 | NLVRDLPQGFSALEP | 53 | RMAGNGCDAALALLL | 53 | TDHSSSSDNIALLVQ |
| 54 | DLPQGFSALEPLVDL | 54 | NGCDAALALLLLDRL | | |

| | | | |
|---|---|---|---|
| 55 | GFSALEPLVDLPIGI | 55 | AALALLLLDRLNQLE |
| 56 | LEPLVDLPIGINITR | 56 | LLLLDRLNQLESKMS |
| 57 | VDLPIGINITRFQTL | 57 | DRLNQLESKMSGKGQ |
| 58 | IGINITRFQTLLALH | 58 | QLESKMSGKGQQQQG |
| 59 | ITRFQTLLALHRSYL | 59 | KMSGKGQQQQGQTVT |
| 60 | QTLLALHRSYLTPGD | 60 | KGQQQQGQTVTKKSA |
| 61 | ALHRSYLTPGDSSSG | 61 | QQGQTVTKKSAAEAS |
| 62 | SYLTPGDSSSGWTAG | 62 | TVTKKSAAEASKKPR |
| 63 | PGDSSSGWTAGAAAY | 63 | KSAAEASKKPRQKRT |
| 64 | SSGWTAGAAAYYVGY | 64 | EASKKPRQKRTATKA |
| 65 | TAGAAAYYVGYLQPR | 65 | KPRQKRTATKAYNVT |
| 66 | AAYYVGYLQPRTFLL | 66 | KRTATKAYNVTQAFG |
| 67 | VGYLQPRTFLLKYNE | 67 | TKAYNVTQAFGRRGP |
| 68 | QPRTFLLKYNENGTI | 68 | NVTQAFGRRGPEQTQ |
| 69 | FLLKYNENGTITDAV | 69 | AFGRRGPEQTQGNFG |
| 70 | YNENGTITDAVDCAL | 70 | RGPEQTQGNFGDQEL |
| 71 | GTITDAVDCALDPLS | 71 | QTQGNFGDQELIRQG |
| 72 | DAVDCALDPLSETKC | 72 | NFGDQELIRQGTDYK |
| 73 | CALDPLSETKCTLKS | 73 | QELIRQGTDYKHWPQ |
| 74 | PLSETKCTLKSFTVE | 74 | RQGTDYKHWPQIAQF |
| 75 | TKCTLKSFTVEKGIY | 75 | DYKHWPQIAQFAPSA |
| 76 | LKSFTVEKGIYQTSN | 76 | WPQIAQFAPSASAFF |
| 77 | TVEKGIYQTSNFRVQ | 77 | AQFAPSASAFFGMSR |
| 78 | GIYQTSNFRVQPTES | 78 | PSASAFFGMSRIGME |
| 79 | TSNFRVQPTESIVRF | 79 | AFFGMSRIGMEVTPS |
| 80 | RVQPTESIVRFPNIT | 80 | MSRIGMEVTPSGTWL |
| 81 | TESIVRFPNITNLCP | 81 | GMEVTPSGTWLTYTG |
| 82 | VRFPNITNLCPFGEV | 82 | TPSGTWLTYTGAIKL |
| 83 | NITNLCPFGEVFNAT | 83 | TWLTYTGAIKLDDKD |
| 84 | LCPFGEVFNATRFAS | 84 | YTGAIKLDDKDPNFK |
| 85 | GEVFNATRFASVYAW | 85 | IKLDDKDPNFKDQVI |
| 86 | NATRFASVYAWNRKR | 86 | DKDPNFKDQVILLNK |

| | | | |
|---|---|---|---|
| 87 | FASVYAWNRKRISNC | 87 | NFKDQVILLNKHIDA |
| 88 | YAWNRKRISNCVADY | 88 | QVILLNKHIDAYKTF |
| 89 | RKRISNCVADYSVLY | 89 | LNKHIDAYKTFPPTE |
| 90 | SNCVADYSVLYNSAS | 90 | IDAYKTFPPTEPKKD |
| 91 | ADYSVLYNSASFSTF | 91 | KTFPPTEPKKDKKKK |
| 92 | VLYNSASFSTFKCYG | 92 | PTEPKKDKKKKAYET |
| 93 | SASFSTFKCYGVSPT | 93 | KKDKKKKAYETQALP |
| 94 | STFKCYGVSPTKLND | 94 | KKKAYETQALPQRQK |
| 95 | CYGVSPTKLNDLCFT | 95 | YETQALPQRQKKQQT |
| 96 | SPTKLNDLCFTNVYA | 96 | ALPQRQKKQQTVTLL |
| 97 | LNDLCFTNVYADSFV | 97 | RQKKQQTVTLLPAAD |
| 98 | CFTNVYADSFVIRGD | 98 | QQTVTLLPAADLDDF |
| 99 | VYADSFVIRGDEVRQ | 99 | TLLPAADLDDFSKQL |
| 100 | SFVIRGDEVRQIAPG | 100 | AADLDDFSKQLQQSM |
| 101 | RGDEVRQIAPGQTGK | 101 | DDFSKQLQQSMSSAD |
| 102 | VRQIAPGQTGKIADY | 102 | KQLQQSMSSADSTQA |
| 103 | APGQTGKIADYNYKL | | |
| 104 | TGKIADYNYKLPDDF | | |
| 105 | ADYNYKLPDDFTGCV | | |
| 106 | YKLPDDFTGCVIAWN | | |
| 107 | DDFTGCVIAWNSNNL | | |
| 108 | GCVIAWNSNNLDSKV | | |
| 109 | AWNSNNLDSKVGGNY | | |
| 110 | NNLDSKVGGNYNYRY | | |
| 111 | SKVGGNYNYRYRLFR | | |
| 112 | GNYNYRYRLFRKSNL | | |
| 113 | YRYRLFRKSNLKPFE | | |
| 114 | LFRKSNLKPFERDIS | | |
| 115 | SNLKPFERDISTEIY | | |
| 116 | PFERDISTEIYQAGS | | |
| 117 | DISTEIYQAGSKPCN | | |
| 118 | EIYQAGSKPCNGVEG | | |

| | |
|---|---|
| 119 | AGSKPCNGVEGFNCY |
| 120 | PCNGVEGFNCYFPLQ |
| 121 | VEGFNCYFPLQSYGF |
| 122 | NCYFPLQSYGFQPTN |
| 123 | PLQSYGFQPTNGVGY |
| 124 | YGFQPTNGVGYQPYR |
| 125 | PTNGVGYQPYRVVVL |
| 126 | VGYQPYRVVVLSFEL |
| 127 | PYRVVVLSFELLHAP |
| 128 | VVLSFELLHAPATVC |
| 129 | FELLHAPATVCGPKK |
| 130 | HAPATVCGPKKSTNL |
| 131 | TVCGPKKSTNLVKNK |
| 132 | PKKSTNLVKNKCVNF |
| 133 | TNLVKNKCVNFNFNG |
| 134 | KNKCVNFNFNGLTGT |
| 135 | VNFNFNGLTGTGVLT |
| 136 | FNGLTGTGVLTESNK |
| 137 | TGTGVLTESNKKFLP |
| 138 | VLTESNKKFLPFQQF |
| 139 | SNKKFLPFQQFGRDI |
| 140 | FLPFQQFGRDIADTT |
| 141 | QQFGRDIADTTDAVR |
| 142 | RDIADTTDAVRDPQT |
| 143 | DTTDAVRDPQTLEIL |
| 144 | AVRDPQTLEILDITP |
| 145 | PQTLEILDITPCSFG |
| 146 | EILDITPCSFGGVSV |
| 147 | ITPCSFGGVSVITPG |
| 148 | SFGGVSVITPGTNTS |
| 149 | VSVITPGTNTSNQVA |
| 150 | TPGTNTSNQVAVLYQ |

| | |
|---|---|
| 151 | NTSNQVAVLYQGVNC |
| 152 | QVAVLYQGVNCTEVP |
| 153 | LYQGVNCTEVPVAIH |
| 154 | VNCTEVPVAIHADQL |
| 155 | EVPVAIHADQLTPTW |
| 156 | AIHADQLTPTWRVYS |
| 157 | DQLTPTWRVYSTGSN |
| 158 | PTWRVYSTGSNVFQT |
| 159 | VYSTGSNVFQTRAGC |
| 160 | GSNVFQTRAGCLIGA |
| 161 | FQTRAGCLIGAEHVN |
| 162 | AGCLIGAEHVNNSYE |
| 163 | IGAEHVNNSYECDIP |
| 164 | HVNNSYECDIPIGAG |
| 165 | SYECDIPIGAGICAS |
| 166 | DIPIGAGICASYQTQ |
| 167 | GAGICASYQTQTNSR |
| 168 | CASYQTQTNSRRRAR |
| 169 | QTQTNSRRRARSVAS |
| 170 | NSRRRARSVASQSII |
| 171 | RARSVASQSIIAYTM |
| 172 | VASQSIIAYTMSLGA |
| 173 | SIIAYTMSLGAENSV |
| 174 | YTMSLGAENSVAYSN |
| 175 | LGAENSVAYSNNSIA |
| 176 | NSVAYSNNSIAIPTN |
| 177 | YSNNSIAIPTNFTIS |
| 178 | SIAIPTNFTISVTTE |
| 179 | PTNFTISVTTEILPV |
| 180 | TISVTTEILPVSMTK |
| 181 | TTEILPVSMTKTSVD |
| 182 | LPVSMTKTSVDCTMY |

| | |
|---|---|
| 183 | MTKTSVDCTMYICGD |
| 184 | SVDCTMYICGDSTEC |
| 185 | TMYICGDSTECSNLL |
| 186 | CGDSTECSNLLLQYG |
| 187 | TECSNLLLQYGSFCT |
| 188 | NLLLQYGSFCTQLNR |
| 189 | QYGSFCTQLNRALTG |
| 190 | FCTQLNRALTGIAVE |
| 191 | LNRALTGIAVEQDKN |
| 192 | LTGIAVEQDKNTQEV |
| 193 | AVEQDKNTQEVFAQV |
| 194 | DKNTQEVFAQVKQIY |
| 195 | QEVFAQVKQIYKTPP |
| 196 | AQVKQIYKTPPIKDF |
| 197 | QIYKTPPIKDFGGFN |
| 198 | TPPIKDFGGFNFSQI |
| 199 | KDFGGFNFSQILPDP |
| 200 | GFNFSQILPDPSKPS |
| 201 | SQILPDPSKPSKRSF |
| 202 | PDPSKPSKRSFIEDL |
| 203 | KPSKRSFIEDLLFNK |
| 204 | RSFIEDLLFNKVTLA |
| 205 | EDLLFNKVTLADAGF |
| 206 | FNKVTLADAGFIKQY |
| 207 | TLADAGFIKQYGDCL |
| 208 | AGFIKQYGDCLGDIA |
| 209 | KQYGDCLGDIAARDL |
| 210 | DCLGDIAARDLICAQ |
| 211 | DIAARDLICAQKFNG |
| 212 | RDLICAQKFNGLTVL |
| 213 | CAQKFNGLTVLPPLL |
| 214 | FNGLTVLPPLLTDEM |

| | |
|---|---|
| 215 | TVLPPLLTDEMIAQY |
| 216 | PLLTDEMIAQYTSAL |
| 217 | DEMIAQYTSALLAGT |
| 218 | AQYTSALLAGTITSG |
| 219 | SALLAGTITSGWTFG |
| 220 | AGTITSGWTFGAGAA |
| 221 | TSGWTFGAGAALQIP |
| 222 | TFGAGAALQIPFAMQ |
| 223 | GAALQIPFAMQMAYR |
| 224 | QIPFAMQMAYRFNGI |
| 225 | AMQMAYRFNGIGVTQ |
| 226 | AYRFNGIGVTQNVLY |
| 227 | NGIGVTQNVLYENQK |
| 228 | VTQNVLYENQKLIAN |
| 229 | VLYENQKLIANQFNS |
| 230 | NQKLIANQFNSAIGK |
| 231 | IANQFNSAIGKIQDS |
| 232 | FNSAIGKIQDSLSST |
| 233 | IGKIQDSLSSTASAL |
| 234 | QDSLSSTASALGKLQ |
| 235 | SSTASALGKLQNVVN |
| 236 | SALGKLQNVVNQNAQ |
| 237 | KLQNVVNQNAQALNT |
| 238 | VVNQNAQALNTLVKQ |
| 239 | NAQALNTLVKQLSSN |
| 240 | LNTLVKQLSSNFGAI |
| 241 | VKQLSSNFGAISSVL |
| 242 | SSNFGAISSVLNDIL |
| 243 | GAISSVLNDILSRLD |
| 244 | SVLNDILSRLDKVEA |
| 245 | DILSRLDKVEAEVQI |
| 246 | RLDKVEAEVQIDRLI |

| | |
|---|---|
| 247 | VEAEVQIDRLITGRL |
| 248 | VQIDRLITGRLQSLQ |
| 249 | RLITGRLQSLQTYVT |
| 250 | GRLQSLQTYVTQQLI |
| 251 | SLQTYVTQQLIRAAE |
| 252 | YVTQQLIRAAEIRAS |
| 253 | QLIRAAEIRASANLA |

**Supplementary Table 3:** Non-structural antigen peptides used for T cell fluorospot assay.

| | NSP12 | | NSP13 | | NSP7 |
|---|---|---|---|---|---|
| 1 | SADAQSFLNRVCGVS | 1 | AVGACVLCNSQTSLR | 1 | SKMSDVKCTSVVLLS |
| 2 | SFLNRVCGVSAARLT | 2 | VLCNSQTSLRCGACI | 2 | VKCTSVVLLSVLQQL |
| 3 | VCGVSAARLTPCGTG | 3 | QTSLRCGACIRRPFL | 3 | VVLLSVLQQLRVESS |
| 4 | AARLTPCGTGTSTDV | 4 | CGACIRRPFLCCKCC | 4 | VLQQLRVESSSKLWA |
| 5 | PCGTGTSTDVVYRAF | 5 | RRPFLCCKCCYDHVI | 5 | RVESSSKLWAQCVQL |
| 6 | TSTDVVYRAFDIYND | 6 | CCKCCYDHVISTSHK | 6 | SKLWAQCVQLHNDIL |
| 7 | VYRAFDIYNDKVAGF | 7 | YDHVISTSHKLVLSV | 7 | QCVQLHNDILLAKDT |
| 8 | DIYNDKVAGFAKFLK | 8 | STSHKLVLSVNPYVC | 8 | HNDILLAKDTTEAFE |
| 9 | KVAGFAKFLKTNCCR | 9 | LVLSVNPYVCNAPGC | 9 | LAKDTTEAFEKMVSL |
| 10 | AKFLKTNCCRFQEKD | 10 | NPYVCNAPGCDVTDV | 10 | TEAFEKMVSLLSVLL |
| 11 | TNCCRFQEKDEDDNL | 11 | NAPGCDVTDVTQLYL | 11 | KMVSLLSVLLSMQGA |
| 12 | FQEKDEDDNLIDSYF | 12 | DVTDVTQLYLGGMSY | 12 | LSVLLSMQGAVDINK |
| 13 | EDDNLIDSYFVVKRH | 13 | TQLYLGGMSYYCKSH | 13 | SMQGAVDINKLCEEM |
| 14 | IDSYFVVKRHTFSNY | 14 | GGMSYYCKSHKLPIS | 14 | VDINKLCEEMLDNRA |
| 15 | VVKRHTFSNYQHEET | 15 | YCKSHKLPISFPLCA | 15 | NKLCEEMLDNRATLQ |
| 16 | TFSNYQHEETIYNLL | 16 | KLPISFPLCANGQVF | | |
| 17 | QHEETIYNLLKDCPA | 17 | FPLCANGQVFGLYKN | | |
| 18 | IYNLLKDCPAVAKHD | 18 | NGQVFGLYKNTCVGS | | |
| 19 | KDCPAVAKHDFFKFR | 19 | GLYKNTCVGSDNVTD | | |
| 20 | VAKHDFFKFRIDGDM | 20 | TCVGSDNVTDFNAIA | | |
| 21 | FFKFRIDGDMVPHIS | 21 | DNVTDFNAIATCDWT | | |
| 22 | IDGDMVPHISRQRLT | 22 | FNAIATCDWTNAGDY | | |
| 23 | VPHISRQRLTKYTMA | 23 | TCDWTNAGDYILANT | | |
| 24 | RQRLTKYTMADLVYA | 24 | NAGDYILANTCTERL | | |
| 25 | KYTMADLVYALRHFD | 25 | ILANTCTERLKLFAA | | |
| 26 | DLVYALRHFDEGNCD | 26 | CTERLKLFAAETLKA | | |
| 27 | LRHFDEGNCDTLKEI | 27 | KLFAAETLKATEETF | | |
| 28 | EGNCDTLKEILVTYN | 28 | ETLKATEETFKLSYG | | |
| 29 | TLKEILVTYNCCDDD | 29 | TEETFKLSYGIATVR | | |
| 30 | LVTYNCCDDDYFNKK | 30 | KLSYGIATVREVLSD | | |
| 31 | CCDDDYFNKKDWYDF | 31 | IATVREVLSDRELHL | | |
| 32 | YFNKKDWYDFVENPD | 32 | EVLSDRELHLSWEVG | | |
| 33 | DWYDFVENPDILRVY | 33 | RELHLSWEVGKPRPP | | |
| 34 | VENPDILRVYANLGE | 34 | SWEVGKPRPPLNRNY | | |
| 35 | ILRVYANLGERVRQA | 35 | KPRPPLNRNYVFTGY | | |
| 36 | ANLGERVRQALLKTV | 36 | LNRNYVFTGYRVTKN | | |
| 37 | RVRQALLKTVQFCDA | 37 | VFTGYRVTKNSKVQI | | |

| | | | |
|---|---|---|---|
| 38 | LLKTVQFCDAMRNAG | 38 | RVTKNSKVQIGEYTF |
| 39 | QFCDAMRNAGIVGVL | 39 | SKVQIGEYTFEKGDY |
| 40 | MRNAGIVGVLTLDNQ | 40 | GEYTFEKGDYGDAVV |
| 41 | IVGVLTLDNQDLNGN | 41 | EKGDYGDAVVYRGTT |
| 42 | TLDNQDLNGNWYDFG | 42 | GDAVVYRGTTTYKLN |
| 43 | DLNGNWYDFGDFIQT | 43 | YRGTTTYKLNVGDYF |
| 44 | WYDFGDFIQTTPGSG | 44 | TYKLNVGDYFVLTSH |
| 45 | DFIQTTPGSGVPVVD | 45 | VGDYFVLTSHTVMPL |
| 46 | TPGSGVPVVDSYYSL | 46 | VLTSHTVMPLSAPTL |
| 47 | VPVVDSYYSLLMPIL | 47 | TVMPLSAPTLVPQEH |
| 48 | SYYSLLMPILTLTRA | 48 | SAPTLVPQEHYVRIT |
| 49 | LMPILTLTRALTAES | 49 | VPQEHYVRITGLYPT |
| 50 | TLTRALTAESHVDTD | 50 | YVRITGLYPTLNISD |
| 51 | LTAESHVDTDLTKPY | 51 | GLYPTLNISDEFSSN |
| 52 | HVDTDLTKPYIKWDL | 52 | LNISDEFSSNVANYQ |
| 53 | LTKPYIKWDLLKYDF | 53 | EFSSNVANYQKVGMQ |
| 54 | IKWDLLKYDFTEERL | 54 | VANYQKVGMQKYSTL |
| 55 | LKYDFTEERLKLFDR | 55 | KVGMQKYSTLQGPPG |
| 56 | TEERLKLFDRYFKYW | 56 | KYSTLQGPPGTGKSH |
| 57 | KLFDRYFKYWDQTYH | 57 | QGPPGTGKSHFAIGL |
| 58 | YFKYWDQTYHPNCVN | 58 | TGKSHFAIGLALYYP |
| 59 | DQTYHPNCVNCLDDR | 59 | FAIGLALYYPSARIV |
| 60 | PNCVNCLDDRCILHC | 60 | ALYYPSARIVYTACS |
| 61 | CLDDRCILHCANFNV | 61 | SARIVYTACSHAAVD |
| 62 | CILHCANFNVLFSTV | 62 | YTACSHAAVDALCEK |
| 63 | ANFNVLFSTVFPLTS | 63 | HAAVDALCEKALKYL |
| 64 | LFSTVFPLTSFGPLV | 64 | ALCEKALKYLPIDKC |
| 65 | FPLTSFGPLVRKIFV | 65 | ALKYLPIDKCSRIIP |
| 66 | FGPLVRKIFVDGVPF | 66 | PIDKCSRIIPARARV |
| 67 | RKIFVDGVPFVVSTG | 67 | SRIIPARARVECFDK |
| 68 | DGVPFVVSTGYHFRE | 68 | ARARVECFDKFKVNS |
| 69 | VVSTGYHFRELGVVH | 69 | ECFDKFKVNSTLEQY |
| 70 | YHFRELGVVHNQDVN | 70 | FKVNSTLEQYVFCTV |
| 71 | LGVVHNQDVNLHSSR | 71 | TLEQYVFCTVNALPE |
| 72 | NQDVNLHSSRLSFKE | 72 | VFCTVNALPETTADI |
| 73 | LHSSRLSFKELLLYA | 73 | NALPETTADIVVFDE |
| 74 | LSFKELLLYAADPAM | 74 | TTADIVVFDEISMAT |
| 75 | LLLYAADPAMHAASG | 75 | VVFDEISMATNYDLS |
| 76 | ADPAMHAASGNLLLD | 76 | ISMATNYDLSVVNAR |

| | | | |
|---|---|---|---|
| 77 | HAASGNLLLDKRTTC | 77 | NYDLSVVNARLRAKH |
| 78 | NLLLDKRTTCFSVAA | 78 | VVNARLRAKHYVYIG |
| 79 | KRTTCFSVAALTNNV | 79 | LRAKHYVYIGDPAQL |
| 80 | FSVAALTNNVAFQTV | 80 | YVYIGDPAQLPAPRT |
| 81 | LTNNVAFQTVKPGNF | 81 | DPAQLPAPRTLLTKG |
| 82 | AFQTVKPGNFNKDFY | 82 | PAPRTLLTKGTLEPE |
| 83 | KPGNFNKDFYDFAVS | 83 | LLTKGTLEPEYFNSV |
| 84 | NKDFYDFAVSKGFFK | 84 | TLEPEYFNSVCRLMK |
| 85 | DFAVSKGFFKEGSSV | 85 | YFNSVCRLMKTIGPD |
| 86 | KGFFKEGSSVELKHF | 86 | CRLMKTIGPDMFLGT |
| 87 | EGSSVELKHFFFAQD | 87 | TIGPDMFLGTCRRCP |
| 88 | ELKHFFFAQDGNAAI | 88 | MFLGTCRRCPAEIVD |
| 89 | FFAQDGNAAISDYDY | 89 | CRRCPAEIVDTVSAL |
| 90 | GNAAISDYDYYRYNL | 90 | AEIVDTVSALVYDNK |
| 91 | SDYDYYRYNLPTMCD | 91 | TVSALVYDNKLKAHK |
| 92 | YRYNLPTMCDIRQLL | 92 | VYDNKLKAHKDKSAQ |
| 93 | PTMCDIRQLLFVVEV | 93 | LKAHKDKSAQCFKMF |
| 94 | IRQLLFVVEVVDKYF | 94 | DKSAQCFKMFYKGVI |
| 95 | FVVEVVDKYFDCYDG | 95 | CFKMFYKGVITHDVS |
| 96 | VDKYFDCYDGGCINA | 96 | YKGVITHDVSSAINR |
| 97 | DCYDGGCINANQVIV | 97 | THDVSSAINRPQIGV |
| 98 | GCINANQVIVNNLDK | 98 | SAINRPQIGVVREFL |
| 99 | NQVIVNNLDKSAGFP | 99 | PQIGVVREFLTRNPA |
| 100 | NNLDKSAGFPFNKWG | 100 | VREFLTRNPAWRKAV |
| 101 | SAGFPFNKWGKARLY | 101 | TRNPAWRKAVFISPY |
| 102 | FNKWGKARLYYDSMS | 102 | WRKAVFISPYNSQNA |
| 103 | KARLYYDSMSYEDQD | 103 | FISPYNSQNAVASKI |
| 104 | YDSMSYEDQDALFAY | 104 | NSQNAVASKILGLPT |
| 105 | YEDQDALFAYTKRNV | 105 | VASKILGLPTQTVDS |
| 106 | ALFAYTKRNVIPTIT | 106 | LGLPTQTVDSSQGSE |
| 107 | TKRNVIPTITQMNLK | 107 | QTVDSSQGSEYDYVI |
| 108 | IPTITQMNLKYAISA | 108 | SQGSEYDYVIFTQTT |
| 109 | QMNLKYAISAKNRAR | 109 | YDYVIFTQTTETAHS |
| 110 | YAISAKNRARTVAGV | 110 | FTQTTETAHSCNVNR |
| 111 | KNRARTVAGVSICST | 111 | ETAHSCNVNRFNVAI |
| 112 | TVAGVSICSTMTNRQ | 112 | CNVNRFNVAITRAKV |
| 113 | SICSTMTNRQFHQKL | 113 | FNVAITRAKVGILCI |
| 114 | MTNRQFHQKLLKSIA | 114 | TRAKVGILCIMSDRD |
| 115 | FHQKLLKSIAATRGA | 115 | GILCIMSDRDLYDKL |

| | | | |
|---|---|---|---|
| 116 | LKSIAATRGATVVIG | 116 | MSDRDLYDKLQFTSL |
| 117 | ATRGATVVIGTSKFY | 117 | LYDKLQFTSLEIPRR |
| 118 | TVVIGTSKFYGGWHN | 118 | QFTSLEIPRRNVATL |
| 119 | TSKFYGGWHNMLKTV | 119 | FTSLEIPRRNVATLQ |
| 120 | GGWHNMLKTVYSDVE | | |
| 121 | MLKTVYSDVENPHLM | | |
| 122 | YSDVENPHLMGWDYP | | |
| 123 | NPHLMGWDYPKCDRA | | |
| 124 | GWDYPKCDRAMPNML | | |
| 125 | KCDRAMPNMLRIMAS | | |
| 126 | MPNMLRIMASLVLAR | | |
| 127 | RIMASLVLARKHTTC | | |
| 128 | LVLARKHTTCCSLSH | | |
| 129 | KHTTCCSLSHRFYRL | | |
| 130 | CSLSHRFYRLANECA | | |
| 131 | RFYRLANECAQVLSE | | |
| 132 | ANECAQVLSEMVMCG | | |
| 133 | QVLSEMVMCGSSLYV | | |
| 134 | MVMCGSSLYVKPGGT | | |
| 135 | SSLYVKPGGTSSGDA | | |
| 136 | KPGGTSSGDATTAYA | | |
| 137 | SSGDATTAYANSVFN | | |
| 138 | TTAYANSVFNICQAV | | |
| 139 | NSVFNICQAVTANVN | | |
| 140 | ICQAVTANVNALLST | | |
| 141 | TANVNALLSTDGNKI | | |
| 142 | ALLSTDGNKIADKYV | | |
| 143 | DGNKIADKYVRNLQH | | |
| 144 | ADKYVRNLQHRLYEC | | |
| 145 | RNLQHRLYECLYRNR | | |
| 146 | RLYECLYRNRDVDTD | | |
| 147 | LYRNRDVDTDFVNEF | | |
| 148 | DVDTDFVNEFYAYLR | | |
| 149 | FVNEFYAYLRKHFSM | | |
| 150 | YAYLRKHFSMMILSD | | |
| 151 | KHFSMMILSDDAVVC | | |
| 152 | MILSDDAVVCFNSTY | | |
| 153 | DAVVCFNSTYASQGL | | |
| 154 | FNSTYASQGLVASIK | | |

| | |
|---|---|
| 155 | ASQGLVASIKNFKSV |
| 156 | VASIKNFKSVLYYQN |
| 157 | NFKSVLYYQNNVFMS |
| 158 | LYYQNNVFMSEAKCW |
| 159 | NVFMSEAKCWTETDL |
| 160 | EAKCWTETDLTKGPH |
| 161 | TETDLTKGPHEFCSQ |
| 162 | TKGPHEFCSQHTMLV |
| 163 | EFCSQHTMLVKQGDD |
| 164 | HTMLVKQGDDYVYLP |
| 165 | KQGDDYVYLPYPDPS |
| 166 | YVYLPYPDPSRILGA |
| 167 | YPDPSRILGAGCFVD |
| 168 | RILGAGCFVDDIVKT |
| 169 | GCFVDDIVKTDGTLM |
| 170 | DIVKTDGTLMIERFV |
| 171 | DGTLMIERFVSLAID |
| 172 | IERFVSLAIDAYPLT |
| 173 | SLAIDAYPLTKHPNQ |
| 174 | AYPLTKHPNQEYADV |
| 175 | KHPNQEYADVFHLYL |
| 176 | EYADVFHLYLQYIRK |
| 177 | FHLYLQYIRKLHDEL |
| 178 | QYIRKLHDELTGHML |
| 179 | LHDELTGHMLDMYSV |
| 180 | TGHMLDMYSVMLTND |
| 181 | DMYSVMLTNDNTSRY |
| 182 | MLTNDNTSRYWEPEF |
| 183 | NTSRYWEPEFYEAMY |
| 184 | WEPEFYEAMYTPHTV |
| 185 | PEFYEAMYTPHTVLQ |

**Protocol**
